# Potential impact of switching from a two- to one-dose gender-neutral routine HPV vaccination program in Canada: A mathematical modeling analysis

**DOI:** 10.1101/2024.05.29.24308112

**Authors:** Mélanie Drolet, Jean-François Laprise, Éléonore Chamberland, Chantal Sauvageau, Sarah Wilson, Gillian H. Lim, Gina Ogilvie, Ashleigh Tuite, Marc Brisson

## Abstract

**Background:** Worldwide, countries are examining whether to implement one-dose HPV vaccination. To inform policy recommendations in Canada, we used mathematical modeling to project the population-level impact and efficiency of switching from two-to one-dose gender-neutral routine HPV vaccination.

**Methods:** We used HPV-ADVISE, an individual-based transmission-dynamic model of HPV infections/diseases, to model 2 provinces (Quebec, Ontario), which represent higher (≈85%) and lower (≈65%) HPV vaccination coverage in Canada. We examined non-inferior and pessimistic scenarios of one-dose efficacy (VE=98%, 90%) and average duration (VD=lifelong, 30 years, 25 years) versus two doses (VE=98%, VD=lifelong). Our main outcomes were the relative reduction in HPV-16 (among females/males) and cervical cancers, and the number of doses needed to prevent one cervical cancer (NNV).

**Results:** Our model projects that one-dose HPV vaccination would avert a similar number of cervical cancers as two doses in Canada, under various non-inferior and pessimistic scenarios. Under the most pessimistic scenario (VD=25 years), one-dose vaccination would avert ∼3 percentage-points fewer cervical cancers than two doses over 100 years. All one-dose scenarios were projected to lead to cervical cancer elimination and were projected to be a substantially more efficient use of vaccine doses compared to two doses (NNVs one-dose vs no vaccination=800-1000; incremental NNVs two-dose vs one-dose vaccination >10,000).

**Interpretation:** If the average duration of one-dose protection is longer than 25 years, individuals would be protected during their peak ages of sexual activity and one-dose vaccination would prevent a similar number of HPV-related cancers, while being a more efficient use of vaccine doses.

## INTRODUCTION

In April 2022, the World Health Organization (WHO) Strategic Advisory Group of Experts on Immunization (SAGE) announced that human papillomavirus (HPV) vaccination with a single dose could be considered for individuals aged 9 to 20 years^1^. This announcement was based on evidence from clinical trials indicating very high and sustained protection against HPV infections with a single dose^2-4^. Since then, updated data from the Costa Rica HPV Vaccine Trial (CVT) showed stable antibodies up to 16 years after one dose vaccination^5^ and the Kenya single-dose HPV-vaccine efficacy randomised-controlled trial (KEN SHE RCT) showed 98% one-dose efficacy after 36 months^6^. The SAGE announcement was also based on modeling suggesting that one-dose vaccination could lead to similar population-level reductions in cervical cancers in low- and middle-income countries (LMICs) as two doses whilst being a more efficient use of vaccine doses, if one-dose duration of protection lasts longer than 20 years^7,8^.

Most of the >35 countries that have adopted one-dose HPV vaccination are LMICs. As of May 2024, about half a dozen high-income countries (HICs) have adopted a one-dose vaccination strategy (e.g. the UK, Australia, Ireland), whilst some have decided to remain with two doses (e.g. Netherland, Spain, Sweden) and many others are examining the issue^9,10^. Several HICs, such as Canada, have been vaccinating against HPV for more than 15 years, initially using three doses (except Quebec) with a switch to two doses in 2015-2016, and are now observing substantial real-world impact of vaccination on HPV infections, precancerous cervical lesions, and cervical cancer^11-14^. Concerns have been raised about possible rebounds in HPV infection or cervical cancer incidence following a switch to one-dose vaccination, if duration of protection of one dose is shorter than two doses. A previous modeling study conducted in the UK suggested that, if one dose leads to much shorter duration of protection than two doses, it could cause a rebound in HPV infections and cervical cancer^15^. However, since the publication of this study, updated data from the final analysis of the KEN SHE RCT^6^ and latest analysis from the CVT^5^ suggests that one-dose vaccine efficacy against HPV infection and duration of protection are higher and longer than many of the scenarios assumed in the modeling study.

Our objective was to examine, using mathematical modeling and up-to-date efficacy and durability data, the population-level impact and efficiency of switching from two-to one-dose gender-neutral routine HPV vaccination in Canada, under non-inferior and pessimistic assumptions of one dose efficacy and duration of protection, for different HPV-related outcomes among females and males (HPV infection, cervical cancer, and other HPV-related cancers). The study was performed to help inform recommendations of the Canadian National Advisory Committee on Immunization (NACI) and the Comité sur l’immunisation du Québec (CIQ).

## METHODS

### Model description

We used HPV-ADVISE (Agent-based Dynamic model for VaccInation and Screening Evaluation), an individual-based transmission-dynamic model, which has been extensively peer reviewed^16-19^, validated to post-vaccination data and to other models^19-22^, and used to inform HPV vaccination policy decisions in Canada and globally^23-28^ (see the technical appendix for an in-depth description of the model: http://www.marc-brisson.net/HPVadvise.pdf). Briefly, HPV-ADVISE simulates HPV transmission through sexual activity, and type-specific progression from infection to cancers of the cervix, anus, oropharynx, vagina, vulva, and penis. Eighteen HPV types are modeled individually. The model can capture a wide variety of HPV vaccination and HPV/cervical cancer screening strategies. For model predictions, we used HPV-ADVISE Canada, which was calibrated to highly stratified Canadian sexual behavior, HPV epidemiology, and cervical screening data (total of 771 targets, data stratified by age and sex, and by HPV type for epidemiological outcomes).

### HPV vaccination coverage and strategies

For model projections, we reproduced historical changes in HPV vaccination programs and coverage by age and gender for Quebec and Ontario from 2007 to 2023 (Appendix Figures A1,A2). These provinces were chosen as they represent a lower and higher bound of vaccination coverage in Canada^29^. In 2007 or 2008, both provinces introduced girls-only routine HPV school-based vaccination with the quadrivalent vaccine and by 2017-18 both had switched to gender-neutral programs with nonavalent vaccine. At the start of HPV vaccination, Quebec had a 5-year catch-up of 14-year-old girls and since the beginning, the vaccine is free of charge up to 17 years old. Furthermore, in 2019/2020, Quebec switched to a 5-year extended schedule with the first dose given at 9 years old and the second dose to be given at 14 years old, from 2024/2025, if required. From 2007/8 to 2023, Quebec had a high mean vaccination coverage (≈85% received two doses by 14 years old) and Ontario had an intermediate vaccination coverage (≈65% received two doses by the end of grade 7).

From 2024 onward, we assumed that vaccination coverage was stable and was not affected by a potential switch to one dose (i.e. 85% coverage in Quebec, and 67% among girls and 62% among boys in Ontario), or change in the program’s target age, in the case of Ontario. To estimate the population-level impact and efficiency of one-dose HPV vaccination, we compared one-dose gender-neutral HPV vaccination of 9-year-olds starting in 2024 to the status quo with two-dose gender-neutral HPV vaccination of 9-year-olds. Of note, given that the focus of the study is on HPV vaccination, all scenarios are with a status quo for cytology-based cervical screening.

### Vaccine efficacy and duration scenarios

Based on results from the India IARC study, the CVT, and the KEN SHE RCT suggesting similar high and sustained protection for one and two doses, our one-dose base case vaccine efficacy and duration of protection scenario assumed non-inferiority of one vs two doses (i.e., 98% lifelong protection against nonavalent HPV types infection). The 98% vaccine efficacy represents the final KEN SHE RCT results^6^. We also examined different pessimistic one-dose scenarios of lower vaccine efficacy (90%) and/or lower average duration of protection (25 or 30 years) (Appendix Table A1). The pessimistic 90% vaccine efficacy scenario represents the lower bound of the one-dose vaccine efficacy 95% confidence interval in the KEN SHE RCT^6^.

The 25 and 30 years of average duration of protection were chosen as pessimistic scenarios given that there is currently no evidence of waning one-dose protection after more than 12 and 16 years of follow-up in the India IARC study and the CVT, respectively^3-5,30^. We modeled duration of protection using a normal distribution, reproducing stable vaccine efficacy for a set number of years. With our pessimistic assumptions, vaccine protection remains stable before dropping rapidly 15-20 years after one-dose vaccination and all vaccinated individuals losing 100% of protection 35-40 years after vaccination (Appendix Figure A3).

### Sensitivity analysis

We performed several sensitivity analyses. First, given that there is greater uncertainty regarding one-dose efficacy for boys as no one-dose RCT exists for males, we examined a scenario where one dose would be non-inferior for girls, but pessimistic for boys (90% vaccine efficacy and 25-year average duration of protection). Second, as currently there is no evidence of waning following one-dose vaccination, there is uncertainty regarding how HPV vaccine protection may wane in the future. We examined an intermediate scenario to our base case where individuals vaccinated with one dose lose 50% of their protection per sexual act as a result of one-dose waning (vs 100% for the base case). Third, given uncertainty related to the number of new sexual partners of mid-adults once one-dose protection could potentially wane, we examined a scenario with lower number of lifetime partners. Finally, we examined a mitigation strategy: switching back to two doses routine vaccination 10 years after the switch to one dose, should ongoing trials show waning of one-dose protection within the next 10 years. The 10-year period was chosen as, in 2034, there would be over 25 years of follow-up in the current one-dose studies (CVT and IARC), which would be sufficient to detect waning if duration of protection is 25 or 30 years.

### Outcomes

To examine the population-level impact of HPV vaccination, our main outcomes were the 1) relative change in HPV-16 infection incidence (among females and males) over time versus no vaccination, 2) relative change in the age-standardized cervical cancer incidence over time versus no vaccination, and 3) percent change in the cumulative incidence of cervical cancer over 100 years (two- or one-dose vaccination vs no vaccination and two-dose vs one-dose vaccination). To examine the efficiency of HPV vaccination, our main outcome was the number of doses needed to prevent one cervical cancer (NNV). This outcome was estimated by dividing the cumulative number of doses given in the population assuming a switch to one dose by the cumulative number of cervical cancers averted over 100 years compared to no vaccination and, incrementally, by dividing the additional number of second doses given by the additional cancers prevented with two-dose vaccination compared to one-dose. Our secondary outcomes were the relative reduction in the incidence of nonavalent high-risk HPV types (16/18/31/33/45/52/58) and all other HPV-related cancers (among females and males) over time versus no vaccination.

For model projections, we identified 50 parameter sets that produced the best fit to data to represent the uncertainty and variability in HPV epidemiology and sexual behavior. Results are presented as the median and 10^th^ and 90^th^ percentiles (80% Uncertainty Interval, “UI”) of model projections for the 50 best fitting parameter sets.

## RESULTS

### Population-level impact of HPV vaccination on HPV-16 infection

The model projects that gender-neutral nonavalent HPV vaccination with two doses or non-inferior one dose would eliminate HPV-16 infections among females and males within the next 15 years in Quebec (vaccination coverage=85%), and reduce HPV-16 infection by >90% in Ontario (vaccination coverage≈65%) (Figure 1, Appendix Tables A2,A3). Under the pessimistic assumption of 90% vaccine efficacy, one-dose vaccination is projected to produce a similar impact on HPV-16 infection compared to two doses in both provinces. Under the most pessimistic scenario of 25-year average duration of protection, one-dose is projected to lead to a 20-25% and 15-20% percentage point rebound in HPV-16 infections among females and males in Quebec and Ontario, respectively. However, when assuming one-dose duration of protection is 30 years, the rebound in infection is about 10% percentage point lower in both provinces. Finally, rebounds in Quebec and Ontario would start more than 25 years (in 2045-2050) after the switch to one dose assuming 25-year average duration of protection, and more than 30 years (in 2050-2055) assuming 30-year average duration of protection.

**Figure 1.**
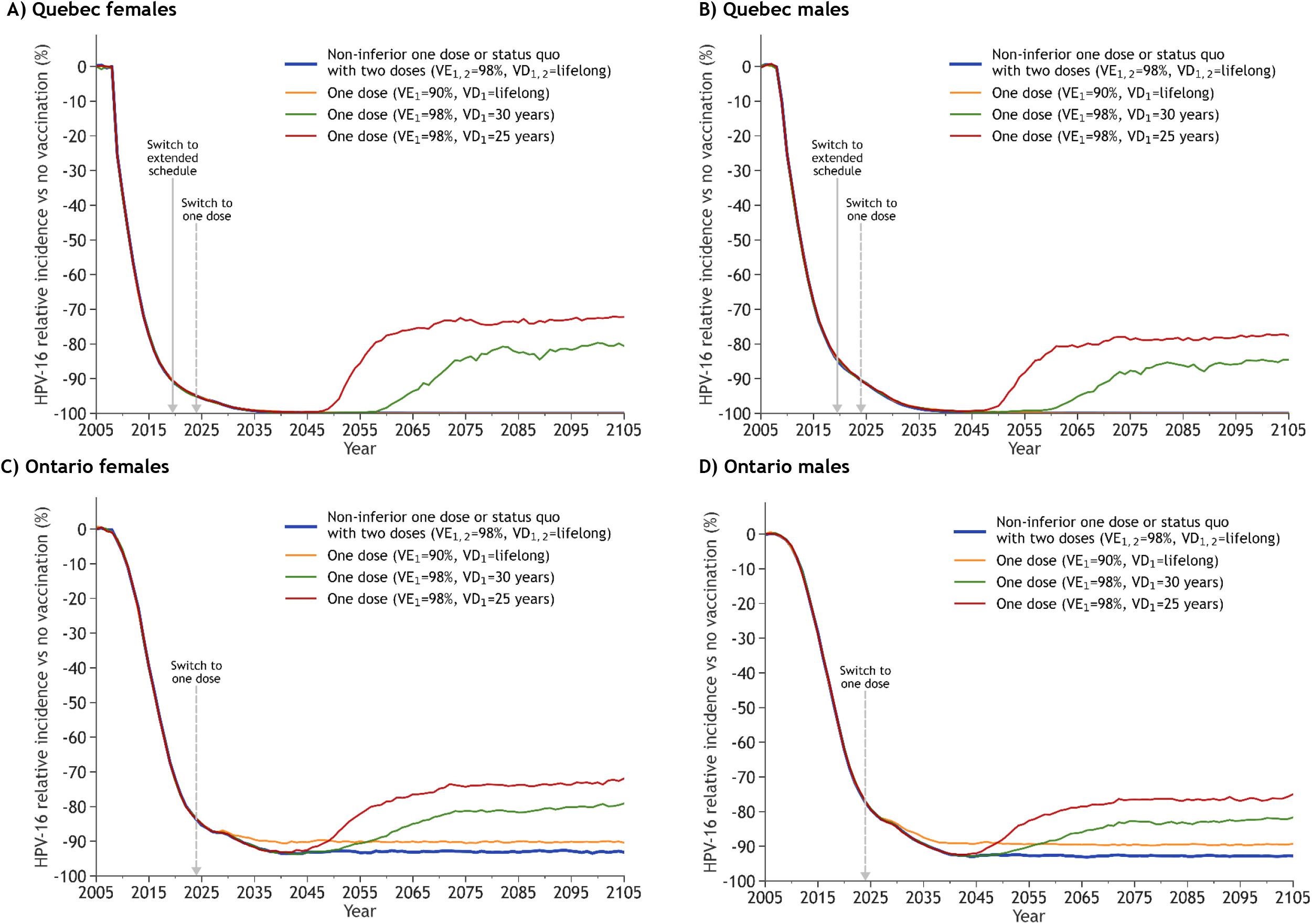
Projected population-level impact of switching to one-dose HPV vaccination for different one-dose efficacy and duration scenarios on HPV-16 incidence among females and males from Quebec and Ontario. VE_*i*_: Vaccine efficacy of dose *i*; VD_*i*_: Vaccine duration of protection of dose *i*. All panels: The lines are the median result of model projections using 50 parameter sets. All scenarios overlap during the first years after the start of vaccination. We chose HPV-16 infection as a main outcome because it contributes most to HPV-related cancers worldwide^43^ and has the highest force of infection. Therefore, it is the hardest to control and would have the highest rebound after a switch to one-dose vaccination, if a rebound happens. Panels A,B: in Quebec the scenarios in blue and yellow lead to the elimination of HPV-16 and are hidden by the x axis. In 2019/2020, Quebec switched to a 5-year extended schedule with the 1^st^ dose given at 9 years old and the 2^nd^ dose to be given at 14 years old, from 2024/2025, if required. If the 2^nd^ dose is not given, Quebec will have switched to one-dose schedule in 2019/2020.

### Population-level impact of HPV vaccination on cervical cancer

The model projects that gender-neutral HPV vaccination with two doses or non-inferior one dose would reduce the age-standardized incidence of cervical cancer by 50% before 2045-2055 and by >85% before the end of the century in both provinces, with a faster and more pronounced reduction in Quebec due to its higher vaccination coverage (Figure 2, Appendix Table A4,A5). Over 100 years, HPV vaccination with two doses or non-inferior one dose is projected to avert about 60% and 55% of all cervical cancers compared to no vaccination in Quebec and Ontario, respectively (Figure 3, Appendix Table A4,A5). Of note, this percentage of averted cancers is estimated among all women since the beginning of vaccination and includes unvaccinated and vaccinated cohorts.

**Figure 2.**
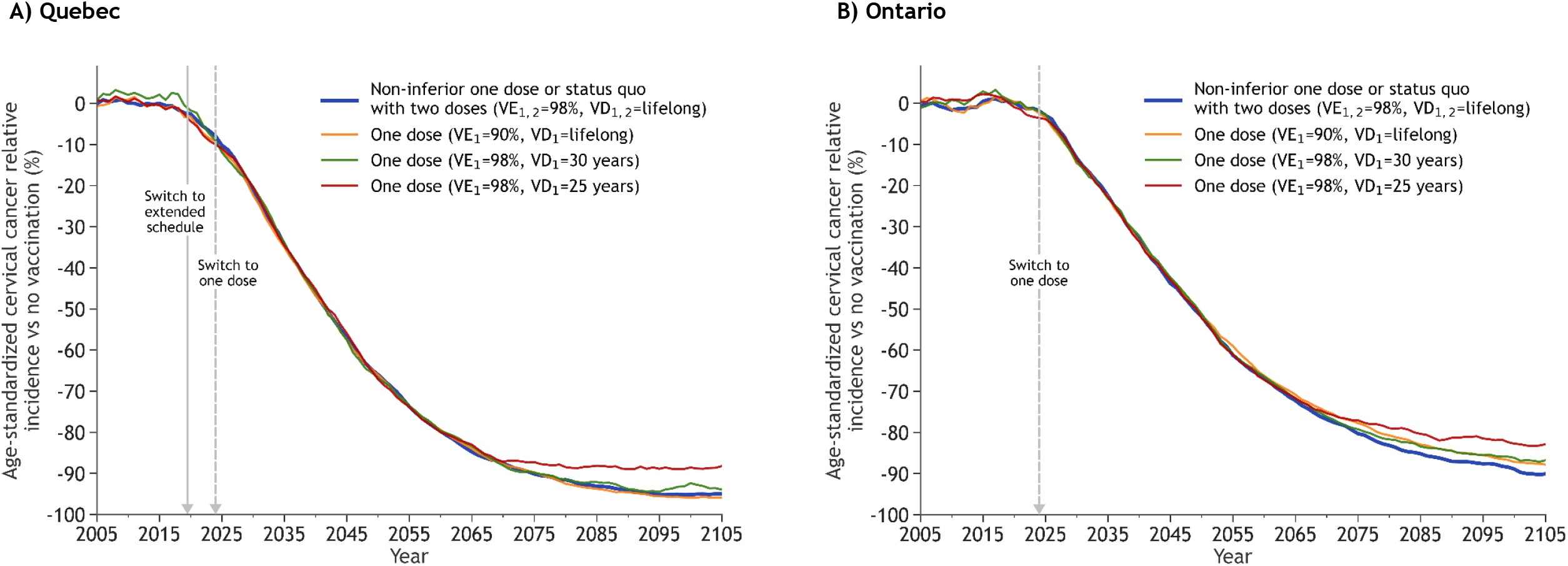
Projected population-level impact of switching to one-dose HPV vaccination for different one-dose efficacy and duration scenarios on cervical cancer incidence in Quebec and Ontario. VE_*i*_: Vaccine efficacy of dose *i*; VD_*i*_: Vaccine duration of protection of dose *i*. All panels: The lines are the median result of model projections using 50 parameter sets. All scenarios overlap during the first years after the start of vaccination. Cervical cancer incidence standardized to the 2015 World population (2017 revision)^44^. Panel A: In 2019/2020, Quebec switched to a 5-year extended schedule with the 1^st^ dose given at 9 years old and the 2^nd^ dose to be given at 14 years old, from 2024/2025, if required. If the 2^nd^ dose is not given, Quebec will have switched to one-dose schedule in 2019/2020.

**Figure 3.**
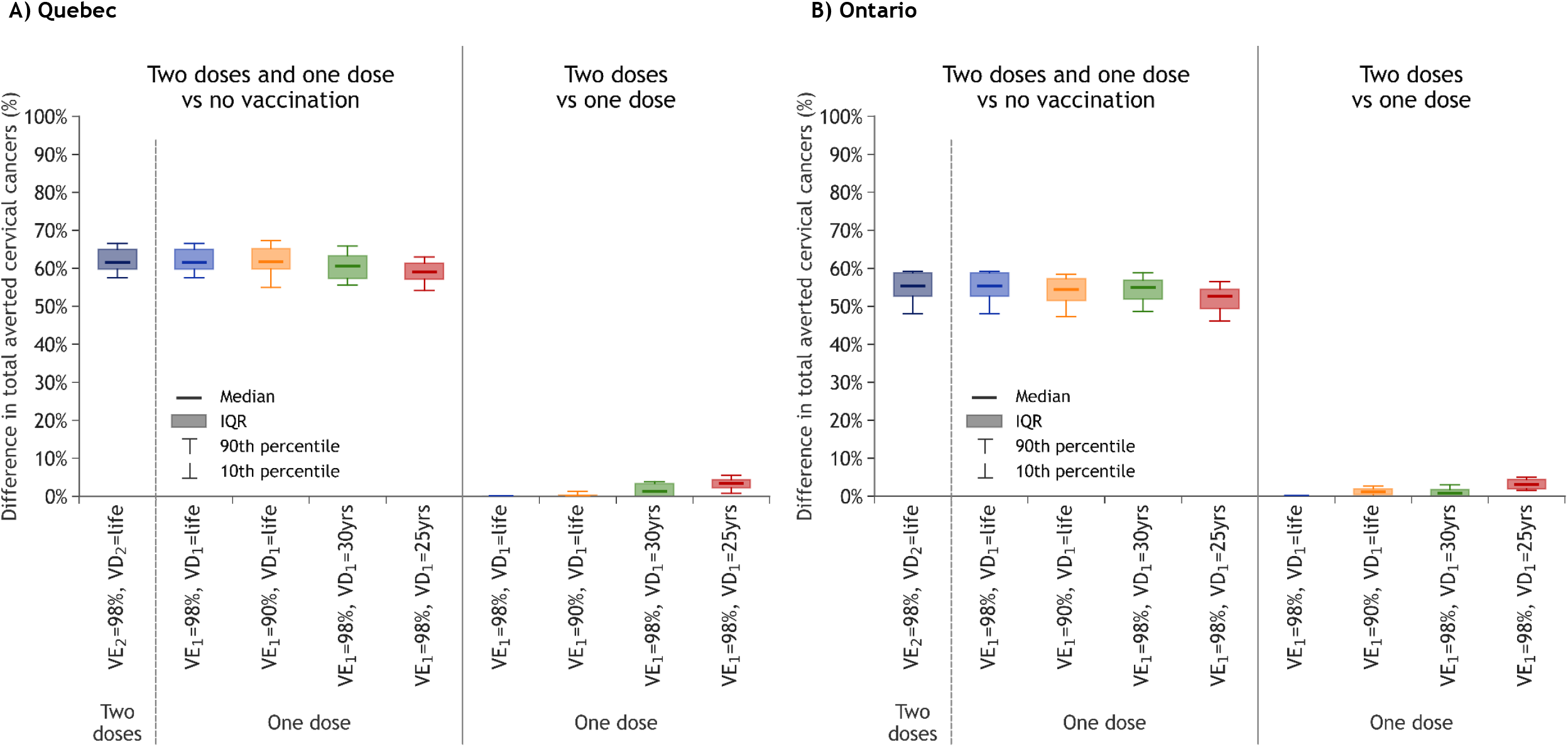
Percent change in the cumulative number of cervical cancers averted over 100 years in Quebec and Ontario for HPV vaccination with two doses and one dose, with different one-dose efficacy and duration scenarios. VE_*i*_: Vaccine efficacy of dose *i*; VD_*i*_: Vaccine duration of protection of dose *i*; IQR: Interquartile range. At the left of each panel, two doses and one dose are compared to no vaccination; at the right of each panel, two doses are compared with one dose, for different one-dose efficacy and duration scenarios. Boxplots represent the median, 10^th^, 25^th^, 75^th^ and 90^th^ percentiles of model projections using 50 parameter sets, illustrating uncertainty related to sexual activity, screening behaviour and the natural history/epidemiology of HPV infection and related diseases. This difference of averted cancer is estimated among all women of all ages since the beginning of vaccination and therefore includes unvaccinated and vaccinated cohorts.

Under the pessimistic one-dose scenarios of 90% vaccine efficacy or 30-year average duration of protection, reductions in cervical cancer over time are projected to be similar to two doses (Figures 2,3, Appendix Tables A4,A5). However, under the most pessimistic scenario of 25-year average duration of protection, one-dose vaccination would avert about 3 percentage-points fewer cervical cancers than two doses over 100 years.

Finally, all one-dose scenarios, including the most pessimistic, are projected to lead to cervical cancer elimination in both provinces, using the WHO elimination threshold (4 cervical cancers/100,000 woman-years) (Appendix Figure A4).

### Efficiency of HPV vaccination

The model projects that one-dose HPV vaccination would be a substantially more efficient use of vaccine doses than two doses, even under the most pessimistic one-dose scenarios (Figure 4, Appendix Tables A4,A5). Compared to no vaccination, the number of doses needed to prevent one cervical cancer (NNV) is projected to be about 800-1000 in Quebec and Ontario for all one-dose scenarios investigated. The incremental NNV for the second dose is about 10,000 for the one-dose pessimistic scenario of 25-year average duration of protection and >25,000 for the other scenarios.

**Figure 4.**
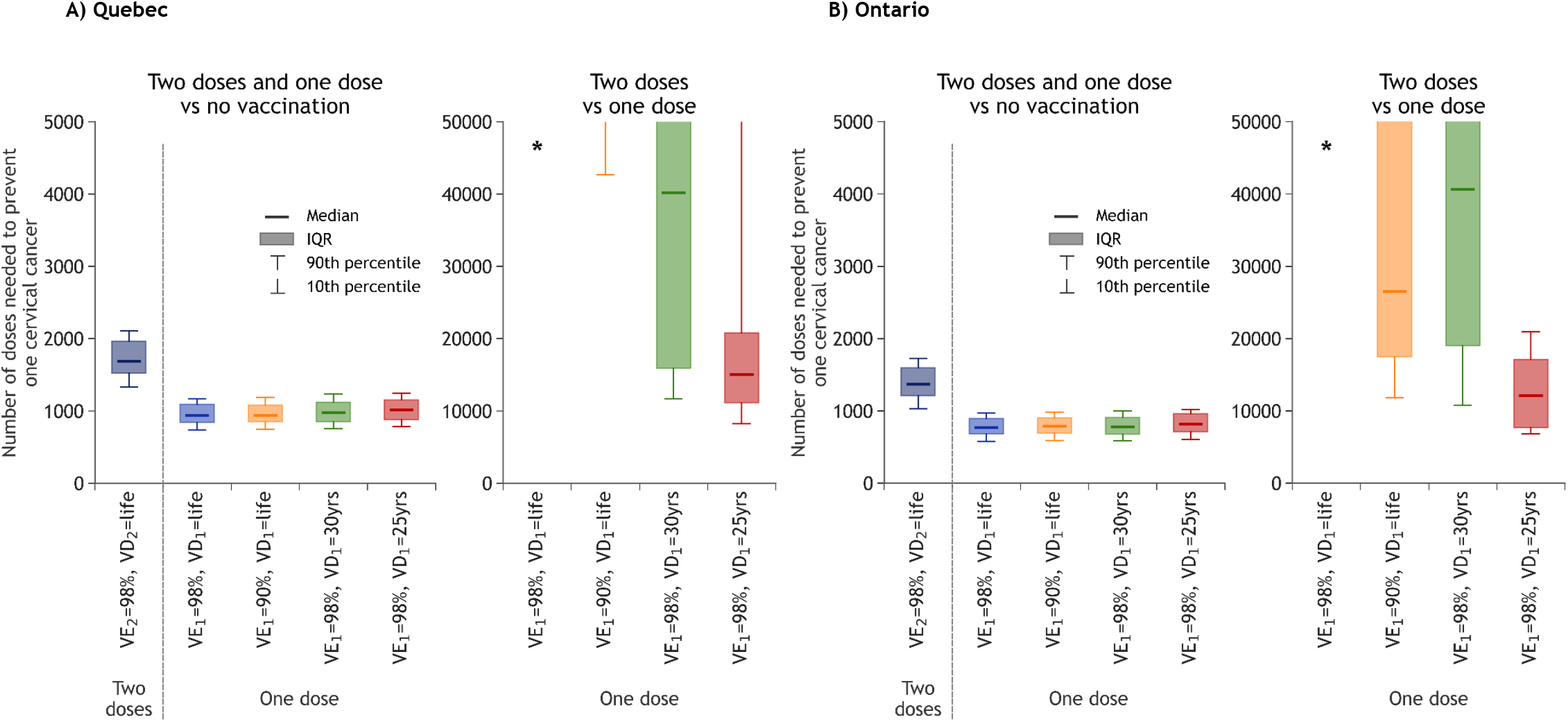
Number of doses needed to prevent one cervical cancer (NNV) in Quebec and Ontario for HPV vaccination with two doses and one dose, with different one-dose efficacy and duration scenarios. VE_*i*_: Vaccine efficacy of dose *i*; VD_*i*_: Vaccine duration of protection of dose *i*. * Model projections of the incremental number of doses needed to prevent one cervical cancer are higher than 50,000 for this scenario. At the left of each panel, two doses and one dose are compared to no vaccination; at the right of each panel, two doses are compared with one dose, for different one-dose efficacy and duration scenarios. Boxplots represent the median, 10^th^, 25^th^, 75^th^ and 90^th^ percentiles of model projections using 50 parameter sets, illustrating uncertainty related to sexual activity, screening behaviour and the natural history/epidemiology of HPV infection and related diseases.

### Secondary outcomes and sensitivity analysis

Our model projects a very limited and delayed rebound in nonavalent high-risk HPV infections and in other HPV-related cancers for the one-dose pessimistic scenarios (vs HPV-16 and cervical cancer, respectively) (Appendix Figures A5,A6).

In sensitivity analysis, assuming pessimistic lower one-dose vaccine efficacy and shorter duration among boys had little to no impact on the projected population-level impact of switching to one-dose vaccination (Appendix Figure A7). Furthermore, if waning leads to partial protection rather than complete loss of protection or if the number of new partners in adults aged >40 years is lower than assumed in our base case, the model projects a smaller rebound in HPV infection and cervical cancer under pessimistic assumptions of one-dose duration, lower NNVs for all one-dose scenarios, and higher incremental NNVs for the second dose (Appendix Figures A8-A10). Finally, under the most pessimistic scenario of an average 25-year duration of protection, switching back to two-dose routine vaccination (after 10 years of one-dose vaccination) would lead to a similar number of cancers averted as remaining with a two-dose strategy (Appendix, Figure A11).

## INTERPRETATION

Our modeling analysis projects that one-dose gender-neutral HPV vaccination would avert a similar number of cervical and other HPV-related cancers as two doses over 100 years in Canada, under a wide range of one-dose vaccine efficacy and duration scenarios. Furthermore, all one-dose vaccination scenarios, even the most pessimistic, were projected to be a substantially more efficient use of vaccine doses compared to two doses and to lead to cervical cancer elimination in Canada within the next 15 to 25 years.

The key remaining uncertainty is the relative durability of one- and two-dose protection. Recent updates of ongoing studies showed sustained efficacy and antibodies up to 12 and 16 years, respectively, after one-dose vaccination^5,30^. Even under the most pessimistic scenario of 25-year average duration of protection, the model projects a limited and delayed rebound in HPV infections, cervical cancer and other HPV-related cancers.

Four key factors can explain this result. First, even with an average duration of protection of 25 years, individuals would be protected during their peak ages of sexual activity and would have, on average, few remaining new partners once efficacy wanes^31-33^. Second, the high vaccination coverage and protection during peak ages of partner acquisition would provide substantial herd effects to unprotected individuals. Third, a potential rebound in HPV infections would start about 25 years following the switch to one-dose vaccination as HPV vaccine-type prevalence is currently very low in Canada (due to high gender-neutral vaccination coverage) and time is needed for the first one-dose cohorts to start losing their protection. Finally, the age at infection would shift from peak ages of sexual activity to the ages when vaccination efficacy would potentially decline. Hence, HPV infection would occur in older individuals leading to a reduced risk of cancer, given the long lag time between infection and development of cancer and fewer life years remaining to develop the cancers.

Our results have important policy implications in Canada, and in other similar HICs examining whether to introduce one-dose HPV vaccination. First, our projections suggest that one-dose vaccination is unlikely to result in a substantial rebound in HPV infection and related cancers in males and females, that the second dose provides small additional population-level benefits in Quebec (vaccination coverage=85%) and Ontario (vaccination coverage≈65%), and that one-dose vaccination would not impact cervical cancer elimination. Under the most pessimistic one-dose duration scenario (25-year average duration of protection), a rebound in HPV-16 infection would occur between 2045-2055. This leaves up to 20 years to detect any significant waning of protection in ongoing studies. If this pessimistic scenario materializes, we have shown that introduction of mitigation strategies, such as reverting to a two-dose strategy after 10 years of one-dose vaccination, would prevent any rebound in cervical cancer. It is therefore important to continue following-up the CVT and IARC studies (which have accrued more than 12 years of follow-up) and to implement HPV surveillance activities in Canada to detect any early signs of increases in prevalence over time. Such surveillance could be facilitated by the introduction of HPV testing in Canadian provinces. Of note, similar concerns about duration of protection were raised when considering a switch from three to two HPV vaccine doses^34^. The decision to switch to two doses was made, at that time, with only four years of two-dose durability data^35^, and modeling results suggesting that two doses would avert as many cancers as three doses if two-dose duration of protection was at least 20 years^36,37^. Second, the COVID-19 pandemic impacted HPV vaccination in Canada, particularly among vulnerable population subgroups (e.g. individuals living in areas with greater socially/materially deprivation or with a greater proportion of immigrants)^38,39^. The economic savings by switching to one-dose vaccination, and its programmatic flexibility, could allow investments to increase vaccination uptake in regions where coverage is suboptimal and in high HPV-burden subgroups to mitigate the pandemic’s impact on programs and to reduce inequalities. Finally, our results are likely generalizable to the other provinces, as Quebec and Ontario cover the range of vaccination coverage in Canada^29^, and to other HICs with similar HPV vaccination coverage, sexual activity (e.g. US, UK, Australia)^40-42^, and HPV epidemiology.

### Limitations

As with all modeling studies, there are limitations related to uncertainty in the data used for projections. We used 50 parameter sets for model projections to capture uncertainty related to sexual activity, screening behaviour and the natural history/epidemiology of HPV infection and related diseases (Figures 3,4). More specifically for one-dose vaccination, three main sources of uncertainty should be considered when using our projections to inform policy decisions: 1) one-dose duration of protection, 2) absence of one-dose efficacy data for boys and other HPV-related cancers, 3) number of new sexual partnerships in adults aged >40 years (once protection would potentially wane). First, we examined wide range of scenarios for one-dose duration, based on the most recent immunogenicity and efficacy data^5,30^. Second, although there is limited one-dose data for boys, our model projected that a pessimistic scenario of combined lower one-dose efficacy (90%) and duration (25-year average duration) for boys would produce a similar population-level impact of one-dose vaccination as a perfect one-dose vaccine for boys, assuming a non-inferior one dose for girls. If gender-neutral vaccination coverage is intermediate/high and vaccine efficacy for girls is high and long lasting, there would be strong enough herd effects to mitigate losses in vaccine efficacy for boys. In addition, even under the most pessimistic one-dose vaccine scenarios, our model projected a more limited and delayed rebound for other HPV-related cancers (vs cervical cancer), given their slower progression from infection to cancer.

Moreover, by preventing HPV infections at the genital sites, one-dose vaccination would also lead to indirect prevention effects to other sites (similar to herd immunity effects). Third, our model slightly over estimates the number of lifetime sexual partners for women over 40 years old compared to recent North American data^31,33^. If this is the case, our projections may overestimate the rebound in HPV infections in scenarios with limited duration of protection. Finally, we did not examine potential changes in screening policies. However, the introduction of more sensitive tests (e.g. HPV testing vs cytology-based screening) or improved participation would reduce the background incidence of cervical cancer, thus further limiting the potential consequences of pessimistic one-dose scenarios and reducing the incremental benefit of a second dose.

## CONCLUSION

Our model projects that one-dose gender-neutral HPV vaccination would avert similar numbers of cervical cancers, HPV infections, and other HPV-related cancers among females and males as two doses in Canada, if vaccine protection remains high during the peak ages of sexual activity. In addition, one-dose vaccination represents a more efficient use of vaccine doses and is projected to lead to cervical cancer elimination in Canada within the next 15 to 25 years. It will be important to continue monitoring one-dose efficacy data over time from the India IARC study and the CVT, which have already cumulated about 15 years of follow-up, to rapidly detect any signs of one-dose efficacy waning. Should we start seeing steep declines in one-dose protection in ongoing studies, policy makers would have sufficient time to introduce mitigation strategies.

## Data Availability

Model outputs are available upon reasonable request.

http://www.marc-brisson.net/HPVadvise.pdf

