## Supplementary Material for "Potential impact of switching from a two- to one-dose gender-neutral routine HPV vaccination program in Canada: A mathematical modeling analysis"

### APPENDIX

**Table A1.** Description of the HPV vaccination scenarios examined.

**Table A2.** Projected population-level impact of switching to one-dose HPV vaccination for different one-dose efficacy and duration scenarios on HPV-16 incidence and on the cumulative number of HPV-16 infections averted over 100 years in Quebec.

**Table A3.** Projected population-level impact of switching to one-dose HPV vaccination for different one-dose efficacy and duration scenarios on HPV-16 incidence and on the cumulative number of HPV-16 infections averted over 100 years in Ontario.

**Table A4.** Projected population-level impact of switching to one-dose HPV vaccination for different one-dose efficacy and duration scenarios on cervical cancer incidence at equilibrium, averted cervical cancers over 100 years and number of doses needed to prevention one cervical cancer in Quebec.

**Table A5.** Projected population-level impact of switching to one-dose HPV vaccination for different one-dose efficacy and duration scenarios on cervical cancer incidence at equilibrium, averted cervical cancers over 100 years and number of doses needed to prevention one cervical cancer in Ontario.

**Figure A1.** Modeled HPV vaccination strategies and coverage in Québec.

**Figure A2.** Modeled HPV vaccination strategies and coverage in Ontario.

**Figure A3.** One-dose vaccine efficacy and immunogenicity over time from clinical trials and estimated by the model for an average duration of 25 years (most pessimistic scenario).

**Figure A4.** Projected population-level impact of switching to one-dose HPV vaccination for different one-dose efficacy and duration scenarios on absolute incidence of cervical cancer in Quebec and Ontario.

**Figure A5.** Projected population-level impact of switching to one-dose HPV vaccination for different one-dose efficacy and duration scenarios on the incidence of nonavalent vaccine high-risk HPV types among females and males from Quebec and Ontario.

**Figure A6.** Projected population-level impact of switching to one-dose HPV vaccination for different one-dose efficacy and duration scenarios on the incidence of all other HPV-related cancers among females and males in Quebec and Ontario.

**Figure A7.** Sensitivity analysis – non-inferior one-dose for girls and worst-case one-dose for boys: projected population-level impact of switching to one-dose on HPV-16 infection and cervical cancer incidence among females and males from Quebec and Ontario.

**Figure A8.** Sensitivity analysis – partial waning and lower sexual activity: projected population-level impact of switching to one-dose on HPV-16 infection and cervical cancer incidence among females and males from Quebec and Ontario.

**Figure A9.** Sensitivity analysis – partial waning and lower sexual activity: percent change in the cumulative number of cervical cancers averted over 100 years in Quebec and Ontario, for HPV vaccination with two doses and one dose, with different one-dose efficacy and duration scenarios.

**Figure A10.** Sensitivity analysis – partial waning and lower sexual activity: number of doses needed to prevent one cervical cancer (NNV) in Quebec and Ontario for HPV vaccination with two doses and one dose, with different one-dose efficacy and duration scenarios.

**Figure A11.** Sensitivity analysis – mitigation strategy: projected population-level impact of switching back to two dose after 10 years of one-dose vaccination on HPV-16 infection and cervical cancer incidence among females and males from Quebec and Ontario.

#### Table A1. Description of the HPV vaccination scenarios examined.

| Vaccination scenarios* | Vaccine efficacy | Mean duration of protection |
|---|---|---|
| **1. Main analysis** | | |
| Comparator: Status quo (current two-dose vaccination strategy & coverage) | 98% | Lifetime |
| Scenarios of switch to one dose : | | |
| • Base-case: Non-inferior one dose | 98% | Lifetime |
| • Pessimistic one-dose efficacy | 90% | Lifetime |
| • Pessimistic one-dose duration | 98% | 30 years |
| • Worst-case one-dose duration | 98% | 25 years |
| • Worst-case one-dose duration and efficacy | 90% | 25 years |
| **2. Sensitivity analysis - non-inferior one-dose for girls & worst-case for boys** | | |
| • Switch to non-inferior one-dose for girls | 98% | Lifetime |
| • Switch to worst-case one-dose for boys | 90% | 25 years |
| **3. Sensitivity analysis - protection waned to a degree of 50%** | | |
| Scenarios of switch to one dose : | | |
| • Base-case: Non-inferior one dose | 98% | Lifetime |
| • Pessimistic one-dose duration | 98% | 30 years |
| • Worst-case one-dose duration | 98% | 25 years |
| **4. Sensitivity analysis - lower level of sexual activity** | | |
| Scenarios of switch to one dose : | | |
| • Base-case: Non-inferior one dose | 98% | Lifetime |
| • Pessimistic one-dose duration | 98% | 30 years |
| • Worst-case one-dose duration | 98% | 25 years |
| **5. Sensitivity analysis - mitigation strategy** | | |
| • Switch to one dose in 2024 worst-case scenario for 10 years | 90% | 25 years |
| • Switch back to routine two doses in 2034 | 98% | Lifetime |

* The vaccination coverage with at least one dose for all scenarios is 85% for girls and boys in Quebec and 67% and 62% for girls and boys, respectively, in Ontario.

**Table A2. Projected population-level impact of switching to one-dose HPV vaccination for different one-dose efficacy and duration scenarios on HPV-16 incidence and on the cumulative number of HPV-16 infections averted over 100 years in Quebec.**

| | QUEBEC | | | | | | | |
|---|---|---|---|---|---|---|---|---|
| | Females | | | | Males | | | |
| | HPV-16 relative incidence at equilibrium (%)* | Percentage point difference in HPV-16 at equilibrium¥ | Time to rebound (years) | Percent change in cumulative HPV-16 infections averted | HPV-16 relative incidence at equilibrium (%)* | Percentage point difference in HPV-16 at equilibrium¥ | Time to rebound (years) | Percent change in cumulative HPV-16 infections averted |
| Scenarios | Median (80% UI) | Median (80% UI) | Median (80% UI) | Median (80% UI) | Median (80% UI) | Median (80% UI) | Median (80% UI) | Median (80% UI) |
| **1. Main analysis** | | | | | | | | |
| **Compared to No vaccination** | | | | | | | | |
| Status quo 2 doses | 100.0 (100.0 - 100.0) | 100.0 (100.0 - 100.0) | NA | 94.7 (94.3 - 94.9) | 100.0 (100.0 - 100.0) | 100.0 (100.0 - 100.0) | NA | 93.1 (91.3 - 93.4) |
| 1 dose non-inferior | 100.0 (100.0 - 100.0) | 100.0 (100.0 - 100.0) | NA | 94.7 (94.3 - 94.9) | 100.0 (100.0 - 100.0) | 100.0 (100.0 - 100.0) | NA | 93.1 (91.3 - 93.4) |
| 1 dose VE=90% | 100.0 (100.0 - 100.0) | 100.0 (100.0 - 100.0) | NA | 94.6 (94.3 - 94.8) | 100.0 (100.0 - 100.0) | 100.0 (100.0 - 100.0) | NA | 93.0 (91.2 - 93.3) |
| 1 dose VD=30 yrs | 80.3 (62.3 - 100.0) | 80.3 (62.3 - 100.0) | 38.0 (31.0 - >100) | 86.2 (79.9 - 94.5) | 84.9 (74.9 - 100.0) | 84.9 (74.9 - 100.0) | 39.0 (31.0 - >100) | 86.6 (83.2 - 92.6) |
| 1 dose VD=25 yrs | 72.6 (52.6 - 81.2) | 72.6 (52.6 - 81.2) | 27.0 (25.9 - 33.0) | 80.8 (72.1 - 86.2) | 77.5 (65.4 - 86.2) | 77.5 (65.4 - 86.2) | 27.0 (26.0 - 35.2) | 81.4 (76.5 - 86.0) |
| 1 dose VE =90%;VD=25 yrs | 71.8 (50.1 - 79.2) | 71.8 (50.1 - 79.2) | 26.0 (23.0 - 30.1) | 79.8 (70.8 - 84.5) | 77.0 (63.2 - 84.8) | 77.0 (63.2 - 84.8) | 26.0 (23.9 - 33.0) | 80.3 (75.2 - 84.8) |
| **Compared to 1 dose** | | | | | | | | |
| 2 vs 1 dose non-inferior | NA | 0.0 (0.0 - 0.0) | NA | 0.0 (0.0 - 0.0) | NA | 0.0 (0.0 - 0.0) | NA | 0.0 (0.0 - 0.0) |
| 2 vs 1 dose VE=90% | NA | 0.0 (0.0 - 0.0) | NA | 0.0 (0.0 - 0.3) | NA | 0.0 (0.0 - 0.0) | NA | 0.1 (-0.1 - 0.4) |
| 2 vs 1 dose VD=30 yrs | NA | 19.7 (0.0 - 37.7) | 38.0 (31.0 - >100) | 8.4 (0.1 - 14.7) | NA | 15.1 (0.0 - 25.1) | 39.0 (31.0 - >100) | 6.5 (0.1 - 10.1) |
| 2 vs 1 dose VD=25 yrs | NA | 27.4 (18.8 - 47.4) | 27.0 (25.0 - 33.0) | 14.1 (8.5 - 22.5) | NA | 22.5 (13.8 - 34.6) | 27.0 (25.9 - 34.3) | 11.7 (5.9 - 16.9) |
| **2. Sensitivity analysis: Non-inferior 1-dose for girls, worst-case for boys** | | | | | | | | |
| **Compared to No vaccination** | | | | | | | | |
| Girls:1 dose non-inferior; Boys: VE=90%;VD=25 yrs | 100.0 (97.5 - 100.0) | 100.0 (97.5 - 100.0) | >100 (33.9 - >100) | 94.5 (93.2 - 94.8) | 100.0 (96.5 - 100.0) | 100.0 (96.5 - 100.0) | >100 (33.9- >100) | 91.9 (91.2 - 93.3) |

| Scenarios | QUEBEC | | | | | | | |
|---|---|---|---|---|---|---|---|---|
| | Females | | | | Males | | | |
| | HPV-16 relative incidence at equilibrium (%)* | Percentage point difference in HPV-16 at equilibrium¥ | Time to rebound (years) | Percent change in cumulative HPV-16 infections averted | HPV-16 relative incidence at equilibrium (%)* | Percentage point difference in HPV-16 at equilibrium¥ | Time to rebound (years) | Percent change in cumulative HPV-16 infections averted |
| | Median (80% UI) | Median (80% UI) | Median (80% UI) | Median (80% UI) | Median (80% UI) | Median (80% UI) | Median (80% UI) | Median (80% UI) |
| **3. Sensitivity analysis: Protection waned to a degree of 50%** | | | | | | | | |
| **Compared to No vaccination** | | | | | | | | |
| 2 doses | 100.0 (100.0 - 100.0) | 100.0 (100.0 - 100.0) | NA | 94.7 (94.3 - 94.9) | 100.0 (100.0 - 100.0) | 100.0 (100.0 - 100.0) | NA | 93.1 (91.3 - 93.4) |
| 1 dose non-inferior | 100.0 (100.0 - 100.0) | 100.0 (100.0 - 100.0) | NA | 94.7 (94.3 - 94.9) | 100.0 (100.0 - 100.0) | 100.0 (100.0 - 100.0) | NA | 93.1 (91.3 - 93.4) |
| 1 dose VD=30 yrs | 92.7 (71.2 - 100.0) | 92.7 (71.2 - 100.0) | 46.0 (32.0 - >100) | 91.6 (83.5 - 94.7) | 95.1 (81.5 - 100.0) | 95.1 (81.5 - 100.0) | 49.0 (34.0 - >100) | 90.3 (85.8 - 93.1) |
| 1 dose VD=25 yrs | 80.4 (64.8 - 89.7) | 80.4 (64.8 - 89.7) | 29.0 (26.9 - 40.2) | 84.7 (77.5 - 90.2) | 84.1 (74.5 - 92.7) | 84.1 (74.5 - 92.7) | 29.5 (27.0 - 42.0) | 84.6 (80.6 - 90.1) |
| **Compared to 1 dose** | | | | | | | | |
| 2 vs 1 dose non-inferior | NA | 0.0 (0.0 - 0.0) | NA | 0.0 (0.0 - 0.0) | NA | 0.0 (0.0 - 0.0) | NA | 0.0 (0.0 - 0.0) |
| 2 vs 1 dose VD=30 yrs | NA | 7.3 (0.0 - 28.8) | 46.0 (32.0 - >100) | 3.1 (-0.1 - 11.1) | NA | 4.9 (0.0 - 18.5) | 49.0 (32.9 - >100) | 1.8 (0.0 - 7.4) |
| 2 vs 1 dose VD=25 yrs | NA | 19.6 (10.3 - 35.2) | 28.5 (26.0 - 38.4) | 9.9 (4.0 - 17.1) | NA | 15.9 (7.3 - 25.5) | 29.0 (26.9 - 42.0) | 8.0 (3.1 - 12.7) |
| **4. Sensitivity analysis: Lower sexual activity** | | | | | | | | |
| **Compared to No vaccination** | | | | | | | | |
| 2 doses | 100.0 (99.0 - 100.0) | 100.0 (99.0 - 100.0) | NA | 94.7 (93.4 - 95.2) | 100.0 (99.3 - 100.0) | 100.0 (99.3 - 100.0) | NA | 92.3 (91.4 - 93.1) |
| 1 dose non-inferior | 100.0 (99.0 - 100.0) | 100.0 (99.0 - 100.0) | NA | 94.7 (93.4 - 95.2) | 100.0 (99.3 - 100.0) | 100.0 (99.3 - 100.0) | NA | 92.3 (91.4 - 93.1) |
| 1 dose VD=30 yrs | 98.6 (94.7 - 100.0) | 98.6 (94.7 - 100.0) | >100 (30.0 - >100) | 93.9 (91.5 - 95.1) | 98.8 (96.2 - 100.0) | 98.8 (96.2 - 100.0) | >100 (44.6 - >100) | 91.6 (90.2 - 93.1) |
| 1 dose VD=25 yrs | 90.0 (83.6 - 97.6) | 90.0 (83.6 - 97.6) | 32.0 (24.0 - 39.0) | 91.2 (86.1 - 92.7) | 91.8 (88.2 - 97.8) | 91.8 (88.2 - 97.8) | 37.0 (28.0 - 46.1) | 89.1 (86.6 - 91.7) |

| | QUEBEC | | | | | | | |
|---|---|---|---|---|---|---|---|---|
| | Females | | | | Males | | | |
| | HPV-16 relative incidence at equilibrium (%)* | Percentage point difference in HPV-16 at equilibrium¥ | Time to rebound (years) | Percent change in cumulative HPV-16 infections averted | HPV-16 relative incidence at equilibrium (%)* | Percentage point difference in HPV-16 at equilibrium¥ | Time to rebound (years) | Percent change in cumulative HPV-16 infections averted |
| Scenarios | Median (80% UI) | Median (80% UI) | Median (80% UI) | Median (80% UI) | Median (80% UI) | Median (80% UI) | Median (80% UI) | Median (80% UI) |
| **Compared to 1 dose** | | | | | | | | |
| 2 vs 1 dose non-inferior | NA | 0.0 (0.0 - 0.0) | NA | 0.0 (0.0 - 0.0) | NA | 0.0 (0.0 - 0.0) | NA | 0.0 (0.0 - 0.0) |
| 2 vs 1 dose VD=30 yrs | NA | 1.3 (0.0 - 4.4) | 69.5 (27.9 - >100) | 0.6 (0.0 - 2.0) | NA | 1.2 (0.0 - 3.8) | >100 (29.0 - >100) | 0.6 (0.0 - 1.3) |
| 2 vs 1 dose VD=25 yrs | NA | 9.9 (2.4 - 15.4) | 30.5 (23.0 - 37.1) | 3.6 (1.5 - 7.3) | NA | 8.1 (2.2 - 11.1) | 33.0 (24.0 - 42.0) | 3.0 (1.4 - 5.0) |
| **5. Sensitivity analysis: Mitigation strategy** | | | | | | | | |
| **Compared to No vaccination** | | | | | | | | |
| Switch back to 2 dose after 10 years of 1 dose VD=25 years | 100.0 (99.4 - 100.0) | 100.0 (99.4 - 100.0) | 27.0 (24.9 - >100) | 93.3 (92.0 - 94.7) | 100.0 (99.3 - 100.0) | 100.0 (99.3 - 100.0) | 27.0 (25.0 - >100) | 91.4 (90.7 - 92.8) |

UI: Uncertainty interval; VE: Vaccine efficacy; VD: Vaccine duration
Results are presented with the median and 80% UI of model projections using 50 parameter sets.
We chose HPV-16 infection as a main outcome because it contributes most to HPV-related cancers worldwide (Guan et al, Human papillomavirus types in 115,789 HPV-positive women: A meta-analysis from cervical infection to cancer, Int J Cancer 2012; 131(10); 2349-59) and has the highest force of infection. Therefore, it is the hardest to control and would have the highest rebound after a switch to one-dose vaccination.
* Relative incidence compared to no vaccination
¥ Percentage point difference of each scenario compared to no vaccination (same results as relative incidence compared to no vaccination) and then compared to 1 dose

**Table A3. Projected population-level impact of switching to one-dose HPV vaccination for different one-dose efficacy and duration scenarios on HPV-16 incidence and on the cumulative number of HPV-16 infections averted over 100 years in Ontario.**

| | ONTARIO | | | | | | | |
|---|---|---|---|---|---|---|---|---|
| | Females | | | | Males | | | |
| | HPV-16 relative incidence at equilibrium (%)* | Percentage point difference in HPV-16 at equilibrium[¥] | Time to rebound (years) | Percent change in cumulative HPV-16 infections averted | HPV-16 relative incidence at equilibrium (%) | Percentage point difference in HPV-16 at equilibrium | Time to rebound (years) | Percent change in cumulative HPV-16 infections averted |
| Scenarios | Median (80% UI) | Median (80% UI) | Median (80% UI) | Median (80% UI) | Median (80% UI) | Median (80% UI) | Median (80% UI) | Median (80% UI) |
| **1. Main analysis** | | | | | | | | |
| **Compared to No vaccination** | | | | | | | | |
| Status quo 2 doses | 92.9 (90.9 - 96.8) | 92.9 (90.9 - 96.8) | NA | 83.3 (80.7 - 86.2) | 92.6 (91.8 - 97.4) | 92.6 (91.8 - 97.4) | NA | 81.6 (79.4 - 83.4) |
| 1 dose non-inferior | 92.9 (90.9 - 96.8) | 92.9 (90.9 - 96.8) | NA | 83.3 (80.7 - 86.2) | 92.6 (91.8 - 97.4) | 92.6 (91.8 - 97.4) | NA | 81.6 (79.4 - 83.4) |
| 1 dose VE=90% | 90.3 (88.2 - 93.3) | 90.3 (88.2 - 93.3) | NA | 81.2 (78.4 - 83.8) | 89.4 (88.6 - 94.7) | 89.4 (88.6 - 94.7) | NA | 79.1 (76.9 - 81.4) |
| 1 dose VD=30 yrs | 79.6 (63.7 - 89.1) | 79.6 (63.7 - 89.1) | 32.0 (27.9 - 38.3) | 77.5 (72.1 - 83.1) | 81.8 (74.0 - 92.2) | 81.8 (74.0 - 92.2) | 34.0 (30.0 - >100) | 76.5 (74.5 - 81.3) |
| 1 dose VD=25 yrs | 72.0 (53.9 - 80.3) | 72.0 (53.9 - 80.3) | 26.0 (24.0 - 30.0) | 73.7 (65.2 - 77.9) | 75.4 (64.8 - 85.0) | 75.4 (64.8 - 85.0) | 27.0 (25.0 - 32.0) | 72.6 (68.5 - 77.3) |
| 1 dose VE =90%;VD=25 yrs | 71.6 (52.9 - 78.9) | 71.6 (52.9 - 78.9) | 25.5 (22.0 - 27.0) | 72.4 (63.9 - 76.3) | 74.3 (62.9 - 83.2) | 74.3 (62.9 - 83.2) | 26.5 (24.0 - 31.0) | 71.2 (66.9 - 75.6) |
| **Compared to 1 dose** | | | | | | | | |
| 2 vs 1 dose non-inferior | NA | 0.0 (0.0 - 0.0) | NA | 0.0 (0.0 - 0.0) | NA | 0.0 (0.0 - 0.0) | NA | 0.0 (0.0 - 0.0) |
| 2 vs 1 dose VE=90% | NA | 3.3 (2.5 - 4.1) | 18.5 (16.0 - >100) | 2.3 (2.0 - 2.6) | NA | 3.4 (2.6 - 3.9) | 23.5 (16.0 - >100) | 2.4 (1.8 - 2.8) |
| 2 vs 1 dose VD=30 yrs | NA | 13.8 (7.3 - 28.4) | 33.0 (29.0 - 41.0) | 5.6 (3.1 - 10.3) | NA | 10.7 (5.2 - 19.2) | 33.0 (29.0 - 48.1) | 4.5 (2.1 - 7.2) |
| 2 vs 1 dose VD=25 yrs | NA | 19.6 (16.6 - 37.9) | 26.0 (21.0 - 28.1) | 9.3 (8.2 - 17.2) | NA | 16.4 (12.3 - 27.8) | 26.0 (21.9 - 29.0) | 8.2 (6.0 - 13.1) |
| **2. Sensitivity analysis: Non-inferior 1-dose for girls, worst-case for boys** | | | | | | | | |
| **Compared to No vaccination** | | | | | | | | |
| Girls:1 dose non-inferior; Boys: VE=90%;VD=25 yrs | 90.3 (87.9 - 94.1) | 90.3 (87.9 - 94.1) | >100 (28.8 - >100) | 81.9 (79.6 - 84.5) | 87.6 (86.0 - 94.4) | 87.6 (86.0 - 94.4) | 41.5 (26.0 - >100) | 78.5 (77.2 - 81.5) |

| Scenarios | ONTARIO | | | | | | | |
|---|---|---|---|---|---|---|---|---|
| | Females | | | | Males | | | |
| | HPV-16 relative incidence at equilibrium (%)* | Percentage point difference in HPV-16 at equilibrium¥ | Time to rebound (years) | Percent change in cumulative HPV-16 infections averted | HPV-16 relative incidence at equilibrium (%) | Percentage point difference in HPV-16 at equilibrium | Time to rebound (years) | Percent change in cumulative HPV-16 infections averted |
| | Median (80% UI) | Median (80% UI) | Median (80% UI) | Median (80% UI) | Median (80% UI) | Median (80% UI) | Median (80% UI) | Median (80% UI) |
| **3. Sensitivity analysis: Protection waned to a degree of 50%** | | | | | | | | |
| **Compared to No vaccination** | | | | | | | | |
| 2 doses | 92.9 (90.9 - 96.8) | 92.9 (90.9 - 96.8) | NA | 83.3 (80.7 - 86.2) | 92.6 (91.8 - 97.4) | 92.6 (91.8 - 97.4) | NA | 81.6 (79.4 - 83.4) |
| 1 dose non-inferior | 92.9 (90.9 - 96.8) | 92.9 (90.9 - 96.8) | NA | 83.3 (80.7 - 86.2) | 92.6 (91.8 - 97.4) | 92.6 (91.8 - 97.4) | NA | 81.6 (79.4 - 83.4) |
| 1 dose VD=30 yrs | 85.3 (71.5 - 91.3) | 85.3 (71.5 - 91.3) | 34.0 (30.0 - 51.4) | 79.9 (74.5 - 83.5) | 86.7 (79.2 - 93.1) | 86.7 (79.2 - 93.1) | 37.5 (30.0 - >100) | 78.5 (76.1 - 81.8) |
| 1 dose VD=25 yrs | 79.2 (63.7 - 84.9) | 79.2 (63.7 - 84.9) | 26.5 (24.0 - 31.0) | 76.3 (69.6 - 79.6) | 80.4 (71.8 - 86.3) | 80.4 (71.8 - 86.3) | 28.0 (25.0 - 34.0) | 75.0 (71.8 - 78.7) |
| **Compared to 1 dose** | | | | | | | | |
| 2 vs 1 dose non-inferior | NA | 0.0 (0.0 - 0.0) | NA | 0.0 (0.0 - 0.0) | NA | 0.0 (0.0 - 0.0) | NA | 0.0 (0.0 - 0.0) |
| 2 vs 1 dose VD=30 yrs | NA | 7.7 (4.6 - 20.2) | 34.0 (29.0 - 43.1) | 3.1 (2.1 - 7.9) | NA | 5.9 (3.4 - 13.8) | 34.5 (30.0 - 64.2) | 2.7 (1.4 - 5.5) |
| 2 vs 1 dose VD=25 yrs | NA | 14.6 (11.4 - 28.2) | 26.0 (21.0 - 29.0) | 7.6 (5.2 - 12.7) | NA | 11.9 (9.9 - 20.9) | 26.5 (21.9 - 30.1) | 5.9 (4.6 - 9.9) |
| **4. Sensitivity analysis: Lower sexual activity** | | | | | | | | |
| **Compared to No vaccination** | | | | | | | | |
| 2 doses | 96.1 (93.7 - 96.4) | 96.1 (93.7 - 96.4) | NA | 86.4 (83.3 - 87.0) | 95.9 (93.9 - 96.3) | 95.9 (93.9 - 96.3) | NA | 83.0 (81.6 - 84.5) |
| 1 dose non-inferior | 96.1 (93.7 - 96.4) | 96.1 (93.7 - 96.4) | NA | 86.4 (83.3 - 87.0) | 95.9 (93.9 - 96.3) | 95.9 (93.9 - 96.3) | NA | 83.0 (81.6 - 84.5) |
| 1 dose VD=30 yrs | 91.3 (88.3 - 92.2) | 91.3 (88.3 - 92.2) | 35.0 (33.0 - 38.1) | 84.6 (81.0 - 85.6) | 91.6 (90.5 - 92.6) | 91.6 (90.5 - 92.6) | 52.0 (35.9 - >100) | 81.6 (79.9 - 83.2) |
| 1 dose VD=25 yrs | 85.3 (79.5 - 88.8) | 85.3 (79.5 - 88.8) | 29.0 (28.0 - 31.0) | 81.8 (76.3 - 82.6) | 86.5 (83.4 - 88.6) | 86.5 (83.4 - 88.6) | 34.0 (30.0 - 37.1) | 79.6 (76.4 - 80.6) |

| Scenarios | ONTARIO | | | | | | | |
|---|---|---|---|---|---|---|---|---|
| | Females | | | | Males | | | |
| | HPV-16 relative incidence at equilibrium (%)* | Percentage point difference in HPV-16 at equilibrium[¥] | Time to rebound (years) | Percent change in cumulative HPV-16 infections averted | HPV-16 relative incidence at equilibrium (%) | Percentage point difference in HPV-16 at equilibrium | Time to rebound (years) | Percent change in cumulative HPV-16 infections averted |
| | Median (80% UI) | Median (80% UI) | Median (80% UI) | Median (80% UI) | Median (80% UI) | Median (80% UI) | Median (80% UI) | Median (80% UI) |
| **Compared to 1 dose** | | | | | | | | |
| 2 vs 1 dose non-inferior | NA | 0.0 (0.0 - 0.0) | NA | 0.0 (0.0 - 0.0) | NA | 0.0 (0.0 - 0.0) | NA | 0.0 (0.0 - 0.0) |
| 2 vs 1 dose VD=30 yrs | NA | 4.8 (3.1 - 5.6) | 33.5 (30.0 - 38.0) | 1.7 (1.3 - 2.3) | NA | 3.7 (2.9 - 4.8) | 36.0 (31.0 - 41.3) | 1.4 (1.2 - 1.8) |
| 2 vs 1 dose VD=25 yrs | NA | 11.0 (5.8 - 14.2) | 26.5 (22.0 - 29.0) | 4.6 (2.8 - 6.9) | NA | 9.4 (5.4 - 10.7) | 27.0 (23.9 - 31.0) | 4.0 (2.5 - 5.1) |
| **5. Sensitivity analysis: Mitigation strategy** | | | | | | | | |
| **Compared to No vaccination** | | | | | | | | |
| Switch back to 2 dose after 10 years of 1 dose VD=25 years | 93.1 (91.2 - 97.0) | 93.1 (91.2 - 97.0) | 29.0 (28.0 - 31.1) | 81.8 (79.4 - 85.0) | 92.7 (91.7 - 97.4) | 92.7 (91.7 - 97.4) | 29.5 (28.0 - >100) | 79.9 (78.3 - 82.5) |

UI: Uncertainty interval; VE: Vaccine efficacy; VD: Vaccine duration
Results are presented with the median and 80% UI of model projections using 50 parameter sets.
We chose HPV-16 infection as a main outcome because it contributes most to HPV-related cancers worldwide (Guan et al, Human papillomavirus types in 115,789 HPV-positive women: A meta-analysis from cervical infection to cancer, Int J Cancer 2012; 131(10); 2349-59) and has the highest force of infection. Therefore, it is the hardest to control and would have the highest rebound after a switch to one-dose vaccination.
* Relative incidence compared to no vaccination.
[¥] Percentage point difference of each scenario compared to no vaccination (same results as relative incidence compared to no vaccination) and then compared to 1 dose.

**Table A4. Projected population-level impact of switching to one-dose HPV vaccination for different one-dose efficacy and duration scenarios on cervical cancer incidence at equilibrium, averted cervical cancers over 100 years and number of doses needed to prevention one cervical cancer in Quebec.**

| Scenarios | QUEBEC | | | | | | |
|---|---|---|---|---|---|---|---|
| | CC relative incidence at equilibrium (%)* | Percentage point difference in CC equilibrium¥ | Time to rebound (years) | Percent change in cumulative CC averted | Absolute difference in cumulative averted CC | Absolute difference in cumulative number of doses | NNV |
| | Median (80% UI) | Median (80% UI) | Median (80% UI) | Median (80% UI) | Median (80% UI) | Median (80% UI) | Median (80% UI) |
| **1. Main analysis** | | | | | | | |
| **Compared to No vaccination** | | | | | | | |
| Status quo 2 doses | 93.2 (86.0 - 99.3) | 93.2 (86.0 - 99.3) | NA | 61.6 (57.4 - 66.6) | 1 469 (1 175 - 1 867) | 2 482 202 (2 478 319 - 2 486 788) | 1 691 (1 330 - 2 111) |
| 1 dose non-inferior | 93.2 (86.0 - 99.3) | 93.2 (86.0 - 99.3) | NA | 61.6 (57.4 - 66.6) | 1 469 (1 175 - 1 867) | 1 375 710 (1 373 658 - 1 377 895) | 937 (737 - 1 170) |
| 1 dose VE=90% | 93.7 (86.2 - 99.0) | 93.7 (86.2 - 99.0) | NA | 61.7 (54.9 - 67.3) | 1 466 (1 159 - 1 838) | 1 375 552 (1 374 356 - 1 378 711) | 939 (749 - 1 186) |
| 1 dose VD=30 yrs | 85.5 (75.6 - 99.1) | 85.5 (75.6 - 99.1) | 75.0 | 60.6 (55.4 - 65.9) | 1 410 (1 115 - 1 829) | 1 375 671 (1 372 238 - 1 379 126) | 977 (754 - 1 234) |
| 1 dose VD=25 yrs | 80.6 (71.0 - 91.8) | 80.6 (71.0 - 91.8) | 61.0 | 59.0 (54.0 - 63.0) | 1 358 (1 100 - 1 759) | 1 375 683 (1 372 891 - 1 379 344) | 1 012 (781 - 1 250) |
| 1 dose VE=90%;VD=25 yrs | 78.2 (69.3 - 89.0) | 78.2 (69.3 - 89.0) | 57.0 | 58.3 (53.1 - 62.9) | 1 348 (1 081 - 1 780) | 1 375 861 (1 372 769 - 1 379 208) | 1 022 (772 - 1 273) |
| **Compared to 1 dose** | | | | | | | |
| 2 vs 1 dose non-inferior | NA | 0.0 (0.0 - 0.0) | NA | 0.0 (0.0 - 0.0) | 0 (0 - 0) | 1 106 578 (1 104 595 - 1 108 880) | >50 000 (>50 000 - >50 000) |
| 2 vs 1 dose VE=90% | NA | 0.0 (-2.2 - 2.2) | NA | -0.2 (-1.4 - 1.3) | 0 (0 - 27) | 1 106 376 (1 103 550 - 1 109 283) | >50 000 (41 599 - >50 000) |
| 2 vs 1 dose VD=30 yrs | NA | 5.1 (-1.4 - 16.3) | 52.0 | 1.3 (-1.1 - 3.8) | 28 (0 - 96) | 1 106 877 (1 101 030 - 1 111 064) | 40 174 (11 599 - >50 000) |
| 2 vs 1 dose VD=25 yrs | NA | 10.9 (5.0 - 23.0) | 51.0 | 3.3 (0.7 - 5.6) | 74 (18 - 134) | 1 107 196 (1 102 062 - 1 109 732) | 15 035 (8 232 - >50 000) |
| **2. Sensitivity analysis: Non-inferior 1-dose for girls, worst-case for boys** | | | | | | | |
| **Compared to No vaccination** | | | | | | | |
| Girls:1 dose non-inferior; Boys: VE=90%;VD=25 yrs | 93.4 (86.2 - 99.7) | 93.4 (86.2 - 99.7) | NA | 62.4 (56.0 - 66.8) | 1 471 (1 180 - 1 819) | 1 375 799 (1 373 291 - 1 377 711) | 935 (756 - 1 167) |

| | QUEBEC | | | | | | |
|---|---|---|---|---|---|---|---|
| | CC relative incidence at equilibrium (%)* | Percentage point difference in CC equilibrium[¥] | Time to rebound (years) | Percent change in cumulative CC averted | Absolute difference in cumulative averted CC | Absolute difference in cumulative number of doses | NNV |
| Scenarios | Median (80% UI) | Median (80% UI) | Median (80% UI) | Median (80% UI) | Median (80% UI) | Median (80% UI) | Median (80% UI) |
| **3. Sensitivity analysis: Protection waned to a degree of 50%** | | | | | | | |
| **Compared to No vaccination** | | | | | | | |
| 2 doses | 93.2 (86.0 - 99.3) | 93.2 (86.0 - 99.3) | NA | 61.6 (57.4 - 66.6) | 1 469 (1 175 - 1 867) | 2 482 202 (2 478 319 - 2 486 788) | 1 691 (1 330 - 2 111) |
| 1 dose non-inferior | 93.2 (86.0 - 99.3) | 93.2 (86.0 - 99.3) | NA | 61.6 (57.4 - 66.6) | 1 469 (1 175 - 1 867) | 1 375 710 (1 373 658 - 1 377 895) | 937 (737 - 1 170) |
| 1 dose VD=30 yrs | 88.7 (77.3 - 98.9) | 88.7 (77.3 - 98.9) | 83.0 | 61.2 (54.3 - 67.2) | 1 450 (1 102 - 1 844) | 1 376 939 (1 373 020 - 1 379 761) | 950 (747 - 1 250) |
| 1 dose VD=25 yrs | 84.3 (77.7 - 98.9) | 84.3 (77.7 - 98.9) | 61.0 | 61.1 (54.5 - 64.4) | 1 420 (1 139 - 1 806) | 1 375 893 (1 372 624 - 1 378 844) | 968 (762 - 1 209) |
| **Compared to 1 dose** | | | | | | | |
| 2 vs 1 dose non-inferior | NA | 0.0 (0.0 - 0.0) | NA | 0.0 (0.0 - 0.0) | 0 (0 - 0) | 1 106 578 (1 104 595 - 1 108 880) | >50 000 (>50 000 - >50 000) |
| 2 vs 1 dose VD=30 yrs | NA | 2.3 (-1.3 - 11.7) | 82.0 | 0.7 (-1.4 - 3.2) | 16 (0 - 75) | 1 106 361 (1 101 650 - 1 109 723) | >50 000 (14 854 - >50 000) |
| 2 vs 1 dose VD=25 yrs | NA | 7.5 (-3.4 - 16.2) | 54.0 | 2.4 (-0.9 - 4.8) | 56 (0 - 130) | 1 106 983 (1 101 531 - 1 111 750) | 19 838 (8 503 - >50 000) |
| **4. Sensitivity analysis: Lower sexual activity** | | | | | | | |
| **Compared to No vaccination** | | | | | | | |
| 2 doses | 95.3 (88.1 - 99.4) | 95.3 (88.1 - 99.4) | NA | 60.7 (57.7 - 63.8) | 1 816 (1 629 - 2 033) | 2 523 526 (2 519 471 - 2 527 693) | 1 389 (1 242 - 1 551) |
| 1 dose non-inferior | 95.3 (88.1 - 99.4) | 95.3 (88.1 - 99.4) | NA | 60.7 (57.7 - 63.8) | 1 816 (1 629 - 2 033) | 1 397 755 (1 395 637 - 1 400 154) | 770 (688 - 859) |
| 1 dose VD=30 yrs | 94.8 (86.7 - 98.1) | 94.8 (86.7 - 98.1) | NA | 60.6 (56.1 - 63.9) | 1 793 (1 631 - 2 105) | 1 398 505 (1 394 836 - 1 401 709) | 779 (665 - 857) |
| 1 dose VD=25 yrs | 92.2 (83.1 - 97.0) | 92.2 (83.1 - 97.0) | 54.0 | 59.7 (54.7 - 64.2) | 1 770 (1 605 - 2 102) | 1 397 257 (1 394 663 - 1 400 703) | 791 (665 - 871) |

| Scenarios | QUEBEC | | | | | | |
|---|---|---|---|---|---|---|---|
| | CC relative incidence at equilibrium (%)* | Percentage point difference in CC equilibrium¥ | Time to rebound (years) | Percent change in cumulative CC averted | Absolute difference in cumulative averted CC | Absolute difference in cumulative number of doses | NNV |
| | Median (80% UI) | Median (80% UI) | Median (80% UI) | Median (80% UI) | Median (80% UI) | Median (80% UI) | Median (80% UI) |
| **Compared to 1 dose** | | | | | | | |
| 2 vs 1 dose non-inferior | NA | 0.0 (0.0 - 0.0) | NA | 0.0 (0.0 - 0.0) | 0 (0 - 0) | 1 125 813 (1 123 991 - 1 127 779) | >50 000 (>50 000 - >50 000) |
| 2 vs 1 dose VD=30 yrs | NA | 0.6 (-1.3 - 4.4) | NA | 0.1 (-1.7 - 1.8) | 3 (0 - 54) | 1 124 810 (1 120 799 - 1 130 776) | >50 000 (20 776 - >50 000) |
| 2 vs 1 dose VD=25 yrs | NA | 3.7 (-1.0 - 6.7) | 72.0 | 0.8 (-1.3 - 3.1) | 25 (0 - 96) | 1 126 483 (1 120 909 - 1 131 305) | 44 942 (11 757 - >50 000) |
| **5. Sensitivity analysis: Mitigation strategy** | | | | | | | |
| **Compared to No vaccination** | | | | | | | |
| Switch back to 2 dose after 10 years of 1 dose VD=25 years | 93.2 (84.9 - 99.2) | 93.2 (84.9 - 99.2) | NA | 61.4 (55.4 - 65.6) | 1 428 (1 198 - 1 824) | 2 283 572 (2 277 826 - 2 289 746) | 1 599 (1 250 - 1 904) |

CC: Cervical cancer; NNV: Number of doses needed to prevent one cervical cancer; UI: Uncertainty interval; VE: Vaccine efficacy; VD: Vaccine duration; Results are presented with the median and 80% UI of model projections using 50 parameter sets.

* Relative incidence compared to no vaccination.

¥ Percentage point difference of each scenario compared to no vaccination (same results as relative incidence compared to no vaccination) and then compared to 1 dose.

**Table A5. Projected population-level impact of switching to one-dose HPV vaccination for different one-dose efficacy and duration scenarios on cervical cancer incidence at equilibrium, averted cervical cancers over 100 years and number of doses needed to prevention one cervical cancer in Ontario.**

| | ONTARIO | | | | | | |
|---|---|---|---|---|---|---|---|
| | CC relative incidence at equilibrium (%)* | Percentage point difference in CC equilibrium[¥] | Time to rebound (years) | Percent change in cumulative CC averted | Absolute difference in cumulative averted CC | Absolute difference in cumulative number of doses | NNV |
| Scenarios | Median (80% UI) | Median (80% UI) | Median (80% UI) | Median (80% UI) | Median (80% UI) | Median (80% UI) | Median (80% UI) |
| **1. Main analysis** | | | | | | | |
| **Compared to No vaccination** | | | | | | | |
| Status quo 2 doses | 90.1 (82.3 - 95.9) | 90.1 (82.3 - 95.9) | NA | 55.4 (47.9 - 59.2) | 1 295 (1 027 - 1 721) | 1 775 084 (1 771 572 - 1 777 780) | 1 370 (1 031 - 1 725) |
| 1 dose non-inferior | 90.1 (82.3 - 95.9) | 90.1 (82.3 - 95.9) | NA | 55.4 (47.9 - 59.2) | 1 295 (1 027 - 1 721) | 994 827 (992 980 - 996 271) | 768 (578 - 967) |
| 1 dose VE=90% | 86.9 (76.2 - 94.9) | 86.9 (76.2 - 94.9) | NA | 54.4 (47.2 - 58.5) | 1 266 (1 008 - 1 690) | 995 046 (992 725 - 996 860) | 788 (588 - 987) |
| 1 dose VD=30 yrs | 82.8 (76.6 - 93.6) | 82.8 (76.6 - 93.6) | 64.0 | 54.9 (48.6 - 58.9) | 1 277 (994 - 1 694) | 994 692 (992 882 - 997 080) | 777 (587 - 1 002) |
| 1 dose VD=25 yrs | 77.0 (67.5 - 88.4) | 77.0 (67.5 - 88.4) | 58.0 | 52.6 (46.0 - 56.5) | 1 216 (977 - 1 660) | 994 643 (993 032 - 996 579) | 819 (599 - 1 019) |
| 1 dose VE=90%;VD=25 yrs | 75.5 (64.6 - 87.3) | 75.5 (64.6 - 87.3) | 65.0 | 51.6 (44.0 - 55.8) | 1 193 (946 - 1 590) | 994 305 (993 274 - 996 461) | 833 (626 - 1 051) |
| **Compared to 1 dose** | | | | | | | |
| 2 vs 1 dose non-inf | NA | 0.0 (0.0 - 0.0) | NA | 0.0 (0.0 - 0.0) | 0 (0 - 0) | 780 208 (778 593 - 781 480) | >50 000 (>50 000 - >50 000) |
| 2 vs 1 dose VE=90% | NA | 3.3 (-1.2 - 7.5) | NA | 1.1 (-0.4 - 2.6) | 29 (0 - 66) | 779 949 (777 192 - 782 423) | 26 519 (11 816 - >50 000) |
| 2 vs 1 dose VD=30 yrs | NA | 4.7 (0.8 - 12.1) | 50.0 | 0.8 (-0.7 - 3.0) | 19 (0 - 72) | 779 573 (777 338 - 782 408) | 40 662 (10 767 - >50 000) |
| 2 vs 1 dose VD=25 yrs | NA | 10.8 (5.6 - 17.7) | 44.0 | 3.1 (1.4 - 5.1) | 64 (37 - 115) | 779 774 (777 432 - 782 631) | 12 112 (6 793 - 21 075) |
| **2. Sensitivity analysis: Non-inferior 1-dose for girls, worst case for boys** | | | | | | | |
| **Compared to No vaccination** | | | | | | | |
| Girls:1 dose non-inferior; Boys: VE=90%;VD=25 yrs | 87.6 (79.4 - 94.7) | 87.6 (79.4 - 94.7) | NA | 54.9 (49.2 - 58.8) | 1 288 (1 026 - 1 700) | 994 874 (993 300 - 996 137) | 771 (585 - 971) |

| Scenarios | ONTARIO | | | | | | |
|---|---|---|---|---|---|---|---|
| | CC relative incidence at equilibrium (%)* | Percentage point difference in CC equilibrium¥ | Time to rebound (years) | Percent change in cumulative CC averted | Absolute difference in cumulative averted CC | Absolute difference in cumulative number of doses | NNV |
| | Median (80% UI) | Median (80% UI) | Median (80% UI) | Median (80% UI) | Median (80% UI) | Median (80% UI) | Median (80% UI) |
| **3. Sensitivity analysis: Protection waned to a degree of 50%** | | | | | | | |
| **Compared to No vaccination** | | | | | | | |
| 2 doses | 90.1 (82.3 - 95.9) | 90.1 (82.3 - 95.9) | NA | 55.4 (47.9 - 59.2) | 1 295 (1 027 - 1 721) | 1 775 084 (1 771 572 - 1 777 780) | 1 370 (1 031 - 1 725) |
| 1 dose non-inferior | 90.1 (82.3 - 95.9) | 90.1 (82.3 - 95.9) | NA | 55.4 (47.9 - 59.2) | 1 295 (1 027 - 1 721) | 994 827 (992 980 - 996 271) | 768 (578 - 967) |
| 1 dose VD=30 yrs | 84.4 (77.7 - 94.2) | 84.4 (77.7 - 94.2) | 73.0 | 54.9 (48.6 - 58.7) | 1 279 (1 021 - 1 685) | 994 974 (992 900 - 997 079) | 779 (591 - 975) |
| 1 dose VD=25 yrs | 81.7 (73.7 - 89.2) | 81.7 (73.7 - 89.2) | 65.0 | 53.9 (47.9 - 57.0) | 1 242 (992 - 1 646) | 994 442 (992 531 - 996 716) | 800 (605 - 1 002) |
| **Compared to 1 dose** | | | | | | | |
| 2 vs 1 dose non-inf | NA | 0.0 (0.0 - 0.0) | NA | 0.0 (0.0 - 0.0) | 0 (0 - 0) | 780 208 (778 593 - 781 480) | >50 000 (>50 000 - >50 000) |
| 2 vs 1 dose VD=30 yrs | NA | 3.6 (-0.4 - 8.8) | 53.0 | 0.7 (-0.7 - 2.1) | 18 (0 - 47) | 779 566 (777 053 - 782 570) | 42 213 (16 659 - >50 000) |
| 2 vs 1 dose VD=25 yrs | NA | 8.6 (2.3 - 13.2) | 44.0 | 2.0 (0.4 - 3.7) | 50 (8 - 81) | 780 082 (777 467 - 782 576) | 15 706 (9 659 - >50 000) |
| **4. Sensitivity analysis: Lower sexual activity** | | | | | | | |
| **Compared to No vaccination** | | | | | | | |
| 2 doses | 92.3 (84.9 - 95.9) | 92.3 (84.9 - 95.9) | NA | 54.0 (51.6 - 58.0) | 1 610 (1 446 - 1 922) | 1 804 084 (1 800 720 - 1 806 617) | 1 121 (940 - 1 249) |
| 1 dose non-inf | 92.3 (84.9 - 95.9) | 92.3 (84.9 - 95.9) | NA | 54.0 (51.6 - 58.0) | 1 610 (1 446 - 1 922) | 1 010 447 (1 008 754 - 1 011 812) | 628 (526 - 700) |
| 1 dose VD=30 yrs | 90.1 (83.3 - 93.9) | 90.1 (83.3 - 93.9) | 83.0 | 54.0 (50.4 - 58.1) | 1 612 (1 465 - 1 902) | 1 010 309 (1 008 811 - 1 011 781) | 627 (532 - 689) |
| 1 dose VD=25 yrs | 87.2 (78.8 - 90.3) | 87.2 (78.8 - 90.3) | 77.0 | 53.5 (49.2 - 57.0) | 1 588 (1 419 - 1 874) | 1 010 360 (1 009 117 - 1 012 360) | 636 (539 - 711) |

| | ONTARIO | | | | | | |
|---|---|---|---|---|---|---|---|
| | CC relative incidence at equilibrium (%)* | Percentage point difference in CC equilibrium[¥] | Time to rebound (years) | Percent change in cumulative CC averted | Absolute difference in cumulative averted CC | Absolute difference in cumulative number of doses | NNV |
| Scenarios | Median (80% UI) | Median (80% UI) | Median (80% UI) | Median (80% UI) | Median (80% UI) | Median (80% UI) | Median (80% UI) |
| **Compared to 1 dose** | | | | | | | |
| 2 vs 1 dose non-inf | NA | 0.0 (0.0 - 0.0) | NA | 0.0 (0.0 - 0.0) | 0 (0 - 0) | 793 563 (791 959 - 794 733) | >50 000 (>50 000 - >50 000) |
| 2 vs 1 dose VD=30 yrs | NA | 2.0 (-0.1 - 4.4) | 54.0 | 0.1 (-1.0 - 1.4) | 3 (0 - 47) | 793 825 (790 860 - 795 781) | >50 000 (16 803 - >50 000) |
| 2 vs 1 dose VD=25 yrs | NA | 5.3 (2.7 - 8.6) | 51.0 | 1.0 (-0.4 - 2.4) | 32 (0 - 80) | 793 367 (790 776 - 795 904) | 24 491 (9 894 - >50 000) |
| **5. Sensitivity analysis: Mitigation strategy** | | | | | | | |
| **Compared to No vaccination** | | | | | | | |
| Switch back to 2 dose after 10 years of 1 dose VD=25 years | 89.6 (80.9 - 95.8) | 89.6 (80.9 - 95.8) | NA | 54.6 (47.6 - 58.8) | 1 265 (1 016 - 1 688) | 1 674 305 (1 671 502 - 1 677 564) | 1 324 (993 - 1 648) |

CC: Cervical cancer; NNV: Number of doses needed to prevent one cervical cancer; UI: Uncertainty interval; VE: Vaccine efficacy; VD: Vaccine duration; Results are presented with the median and 80% UI of model projections using 50 parameter sets.

* Relative incidence compared to no vaccination.

[¥] Percentage point difference of each scenario compared to no vaccination (same results as relative incidence compared to no vaccination) and then compared to 1 dose.

**Figure A1. Modeled HPV vaccination strategies and coverage in Québec.**

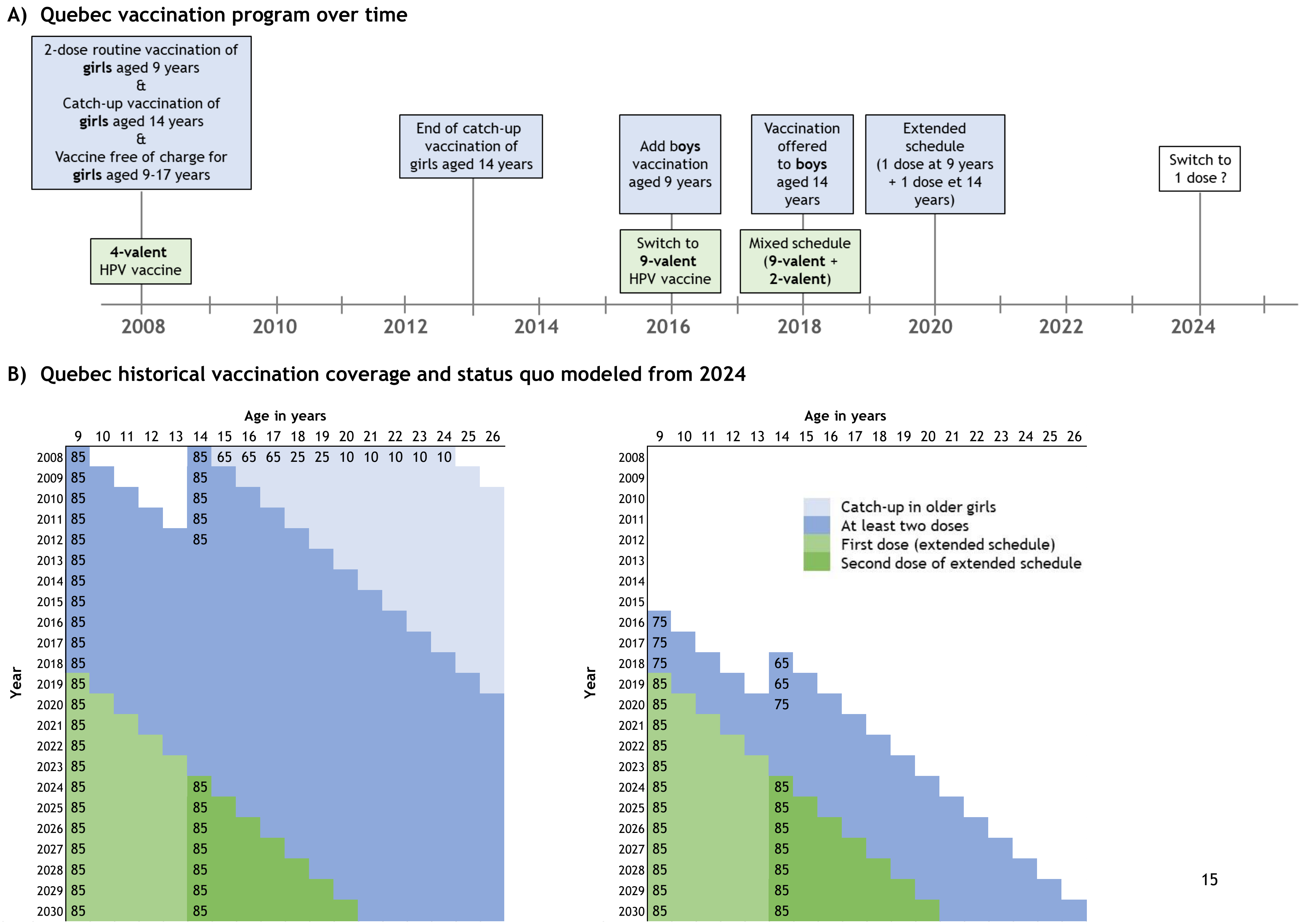

#### C) Quebec historical vaccination coverage and switch to one-dose vaccination modeled from 2024

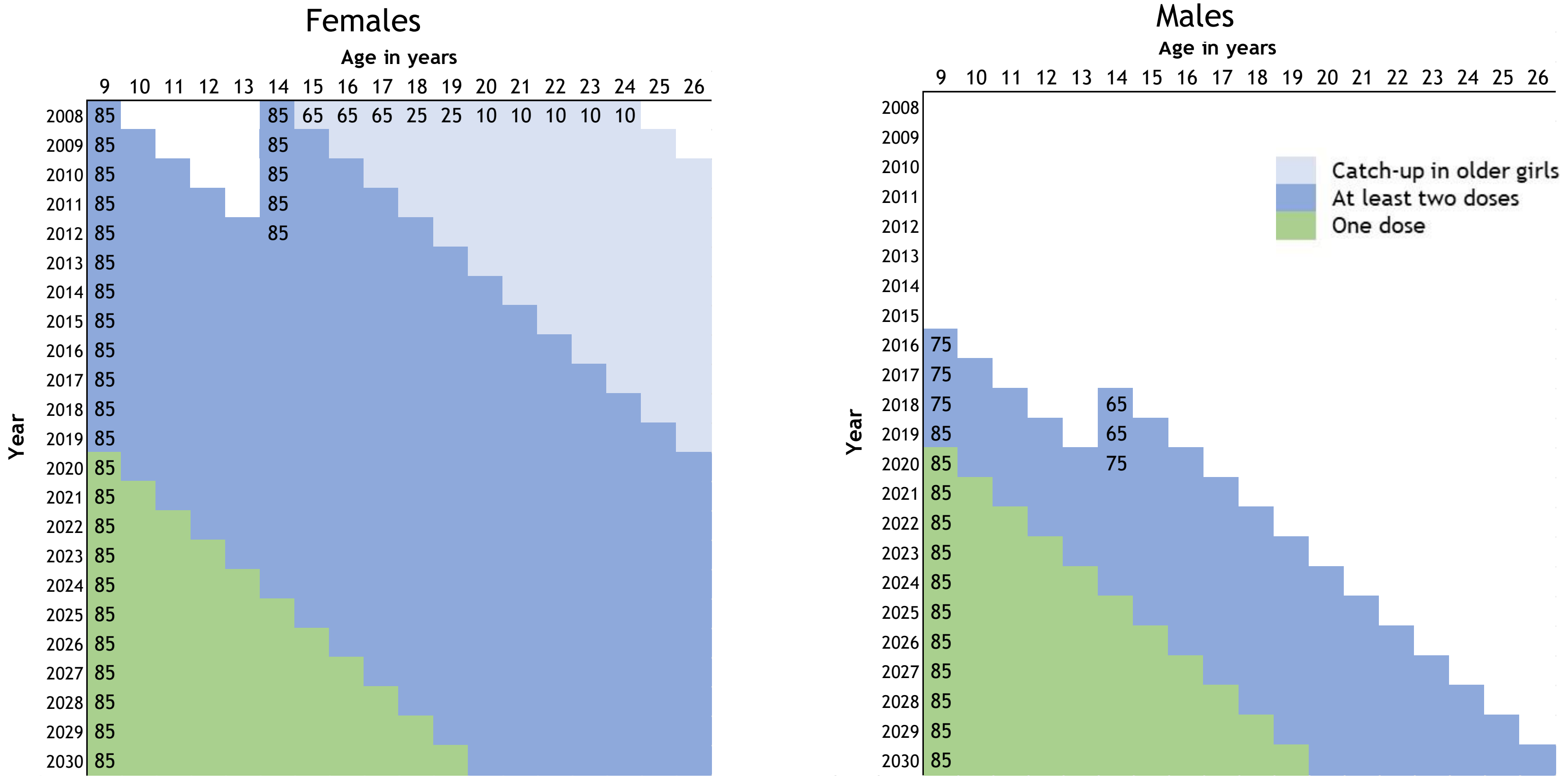


* Coverage as modeled is based on historical coverage available from the Ministère de la Santé et des Services sociaux Québec (MSSS); Coverage values are rounded to nearest 5%

References : Ministère de la Santé et des Services Sociaux du Québec (MSSS). Flash Vigie - Bulletin québécois de vigie, de surveillance et d'intervention en protection de la santé publique - Septembre 2019 Vaccination en milieu scolaire. 2019. https://publications.msss.gouv.qc.ca/msss/fichiers/flashvigie/FlashVigie_vol14_no7.pdf (accessed January 20, 2020); Goggin P, Coutlée F, Defay F, et al. Prévalence des infections au virus du papillome humain (VPH) : résultats de l'étude PIXEL-Portrait de la santé sexuelle des jeunes adultes au Québec, 2013-2014: Institut national de santé publique du Québec (INSPQ), 2015.

**Figure A2. Modeled HPV vaccination strategies and coverage in Ontario.**

A) Ontario vaccination program over time

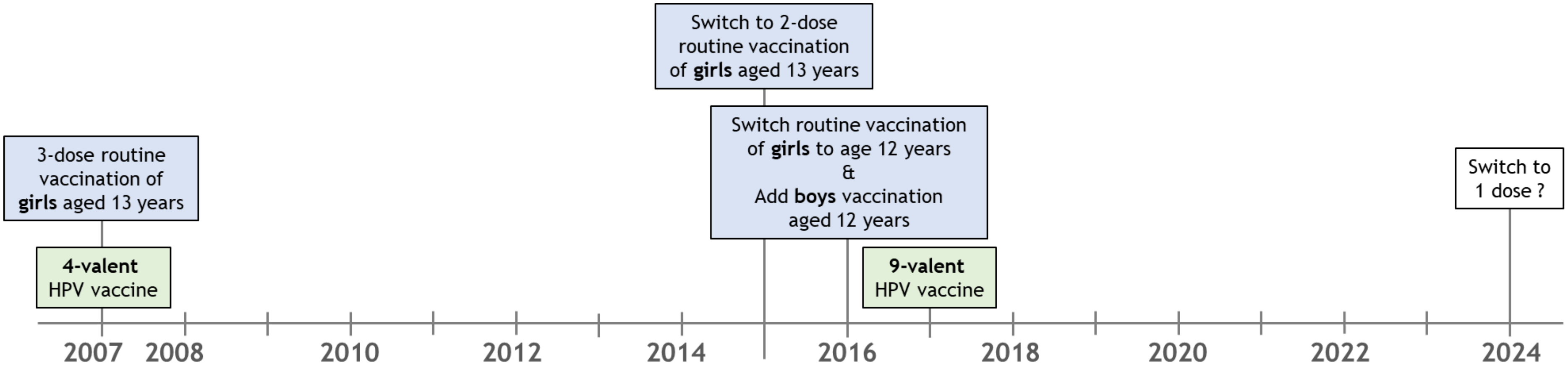


B) Ontario historical vaccination coverage and status quo modeled from 2024*

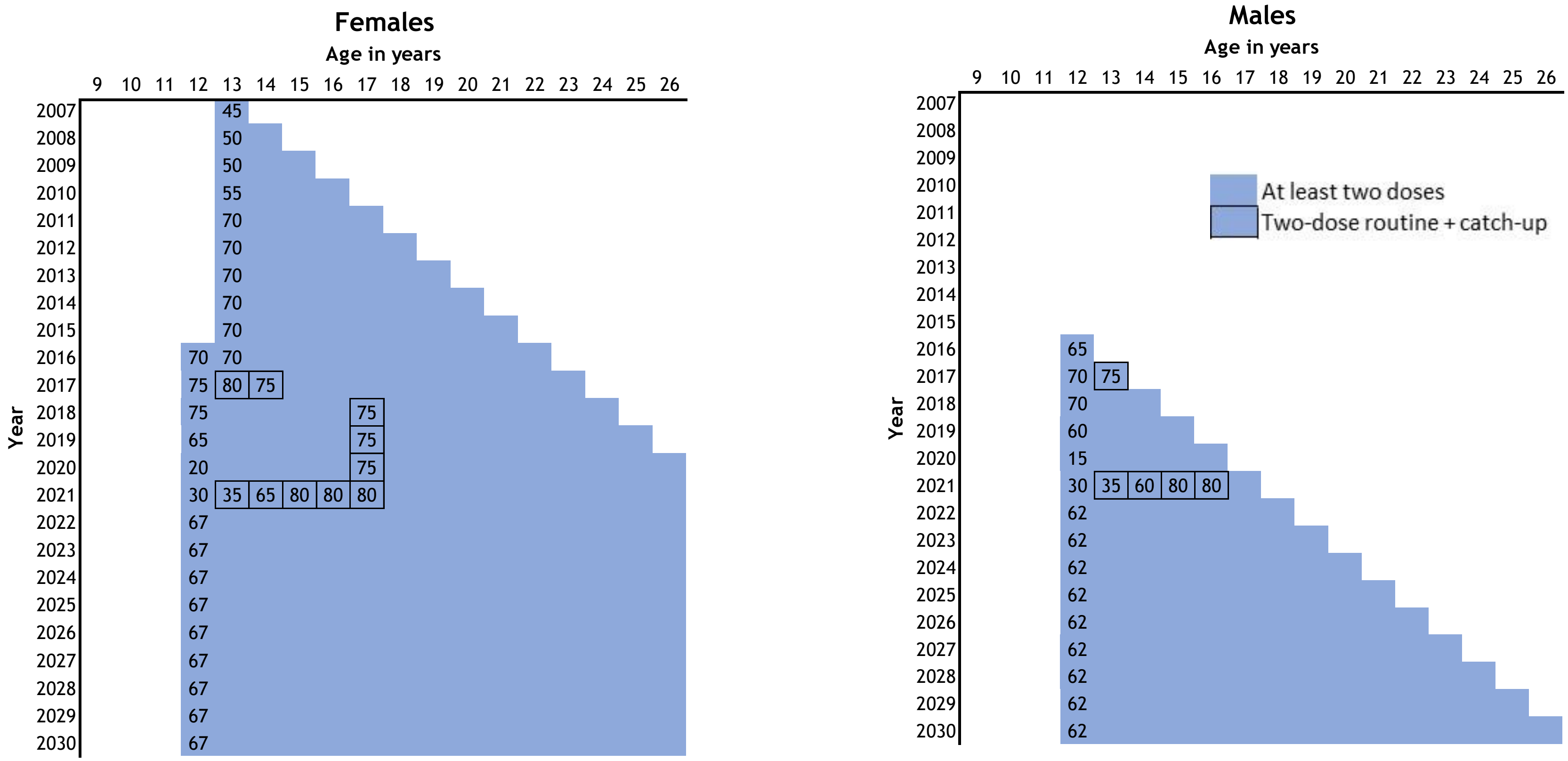

#### C) Ontario historical vaccination coverage and switch to one-dose vaccination modeled from 2024

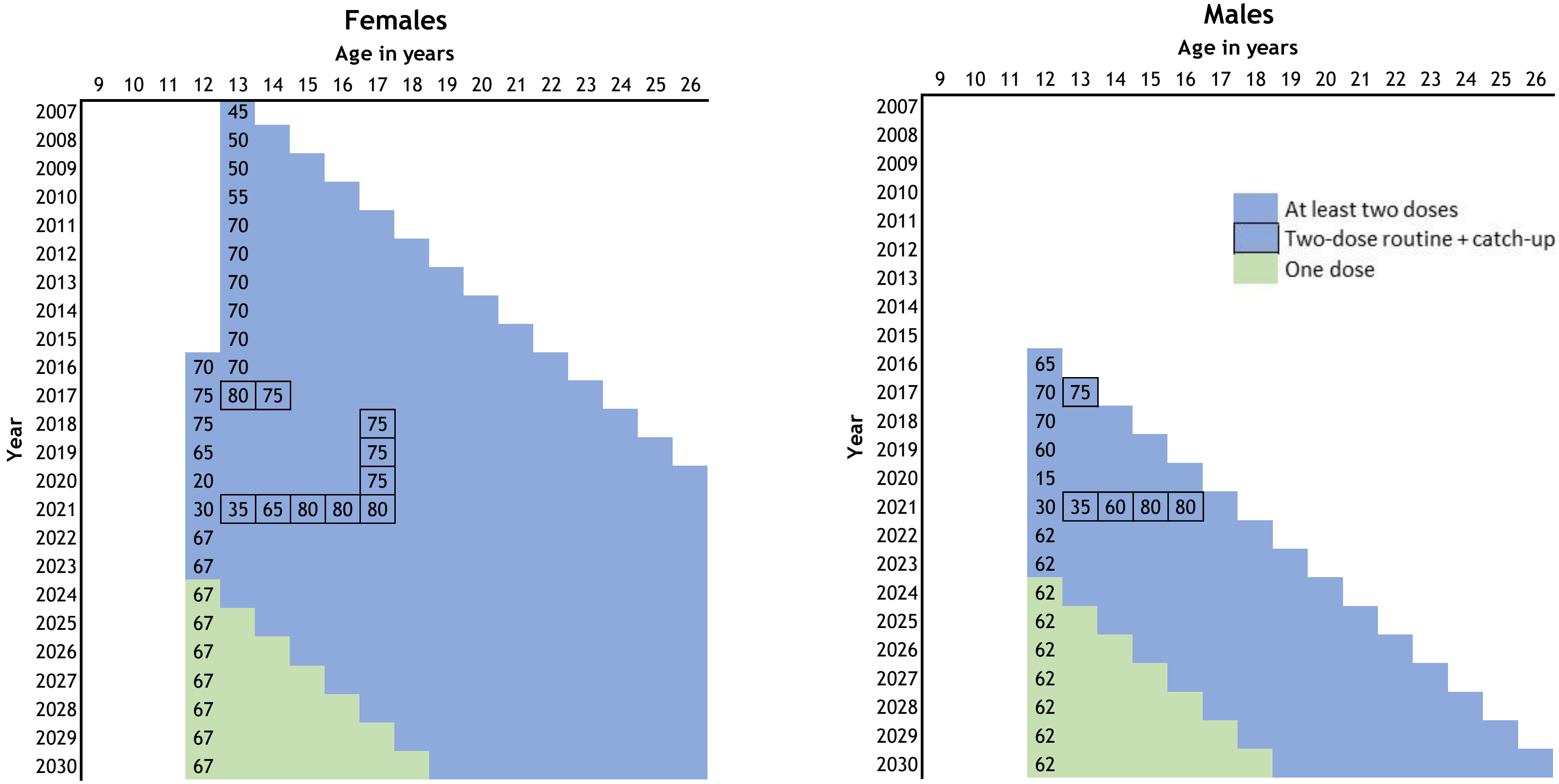


* Coverage as modeled is based on historical coverage available from Pubic Health Ontario
Reference: Public Health Ontario. Immunization Coverage. https://www.publichealthontario.ca/en/health-topics/immunization/vaccine-coverage (accessed April 25, 2024).

**Figure A3.** One-dose vaccine efficacy and immunogenicity over time from clinical trials and estimated by the model for an average duration of 25 years (most pessimistic scenario).

A) Vaccine efficacy

B) Vaccine duration of protection

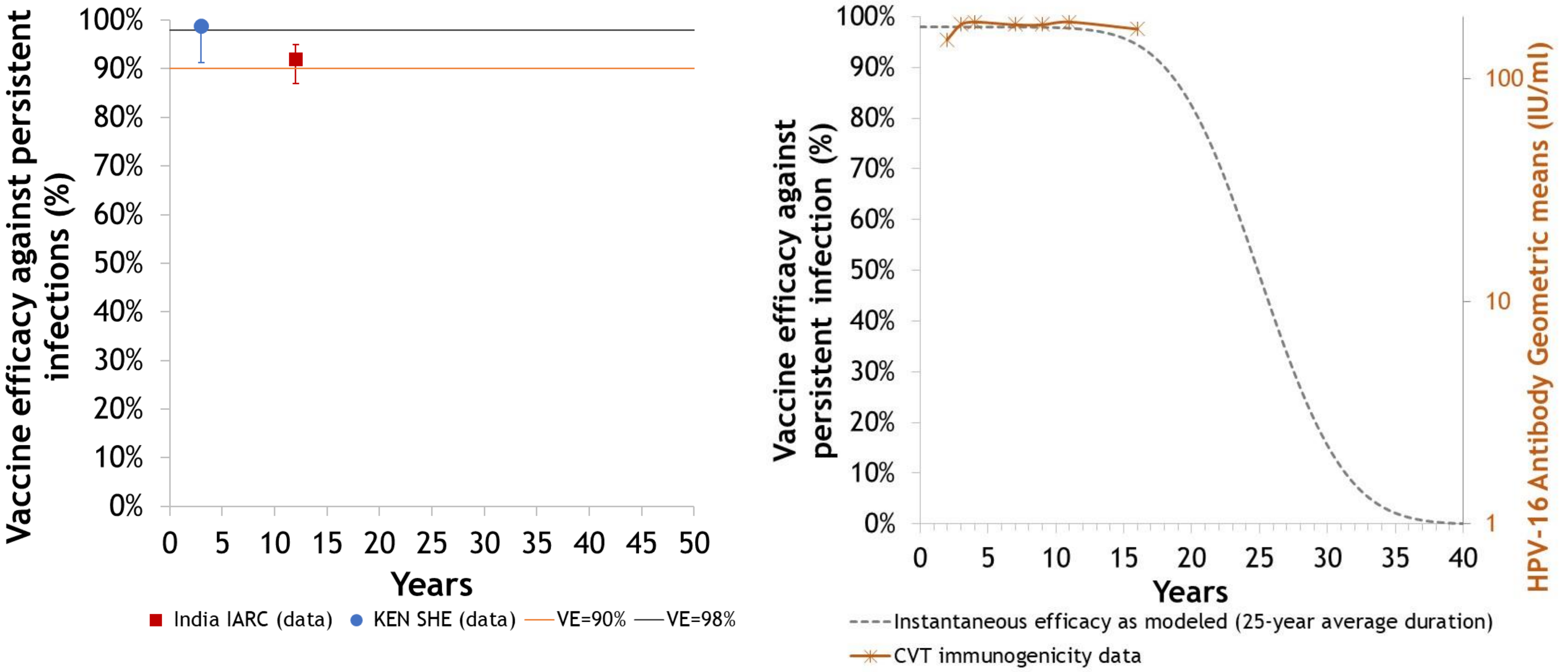


The vaccine efficacy as modeled in HPV-ADVISE represents the instantaneous efficacy

References: **KEN SHE Trial**: Barnabas RV, Brown ER, Onono MA, et al. Durability of single-dose HPV vaccination in young Kenyan women: randomized controlled trial 3-year results. Nature Medicine 2023;29(12):3224-3232. **IARC India study**: Malvi SG, Esmy PO, Muwonge R, *et al*. A prospective cohort study comparing efficacy of one dose of quadrivalent HPV vaccine against two and three doses 15 years post-vaccination Journal National Cancer Institute 2024;Submitted. **Costa Rica Trial (CVT)**: Porras C, Romero B, Kemp T, *et al*. Durability of HPV-16/18 Antibodies 16 years after a Single Dose of the Bivalent HPV Vaccine: The Costa Rica HPV Vaccine Trial (CVT) Journal National Cancer Institute 2024;Submitted.

**Figure A4. Projected population-level impact of switching to one-dose HPV vaccination for different one-dose efficacy and duration scenarios on absolute incidence of cervical cancer in Quebec and Ontario.**

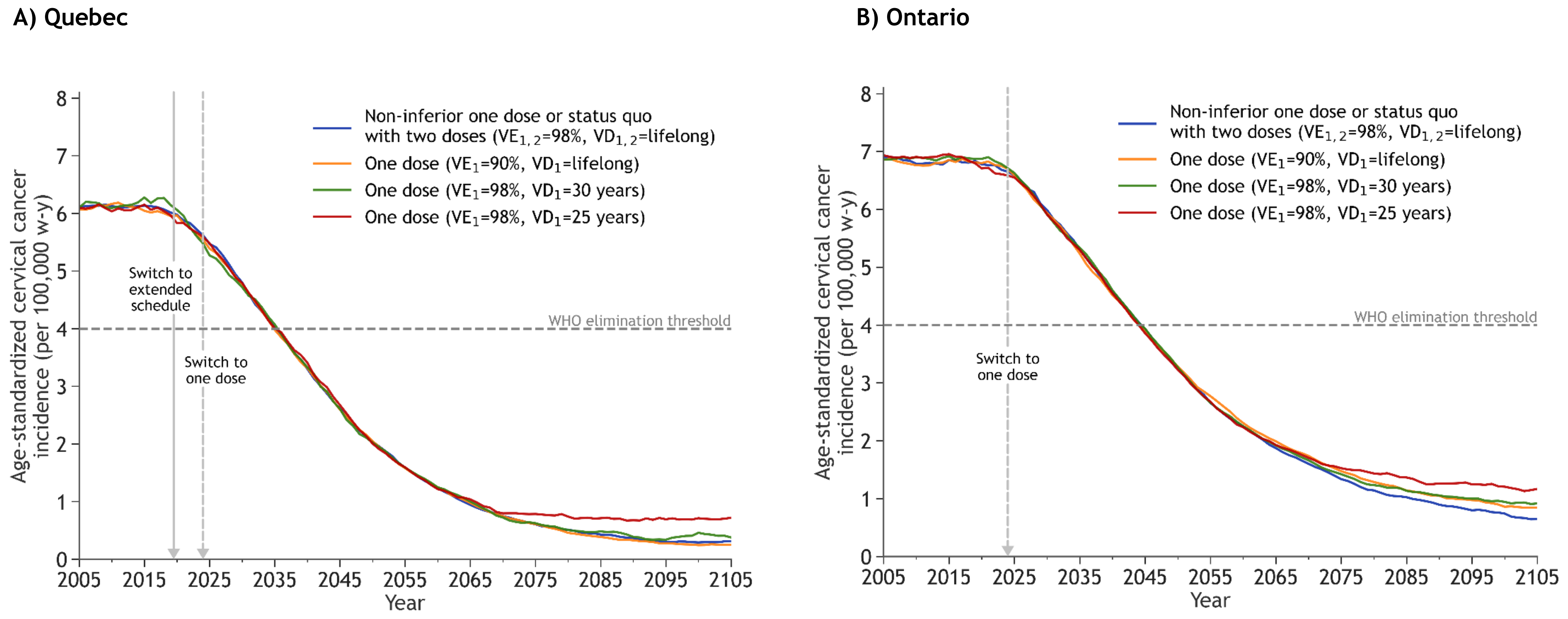


$VE_i$: Vaccine efficacy of dose *i*; $VD_i$: Vaccine duration of protection of dose *i*.

All panels: The lines are the median result of model projections using 50 parameter sets. All scenarios overlap during the first years after the start of vaccination. Cervical cancer incidence standardized to the 2015 World population (2017 revision - United Nations, Department of Economic and Social Affairs, Division P. World Population Prospects: The 2017 Revision, custom data acquired via website. https://esa.un.org/unpd/wpp/dataquery/ )

Panel A: In 2019/2020, Quebec switched to a 5-year extended schedule with the 1st dose given at 9 years old and the 2nd dose to be given at 14 years old, from 2024/2025, if required. If the 2nd dose is not given, Quebec will have switched to one-dose schedule in 2019/2020.

**Figure A5. Projected population-level impact of switching to one-dose HPV vaccination for different one-dose efficacy and duration scenarios on the incidence of nonavalent vaccine high-risk HPV types among females and males from Quebec and Ontario.**

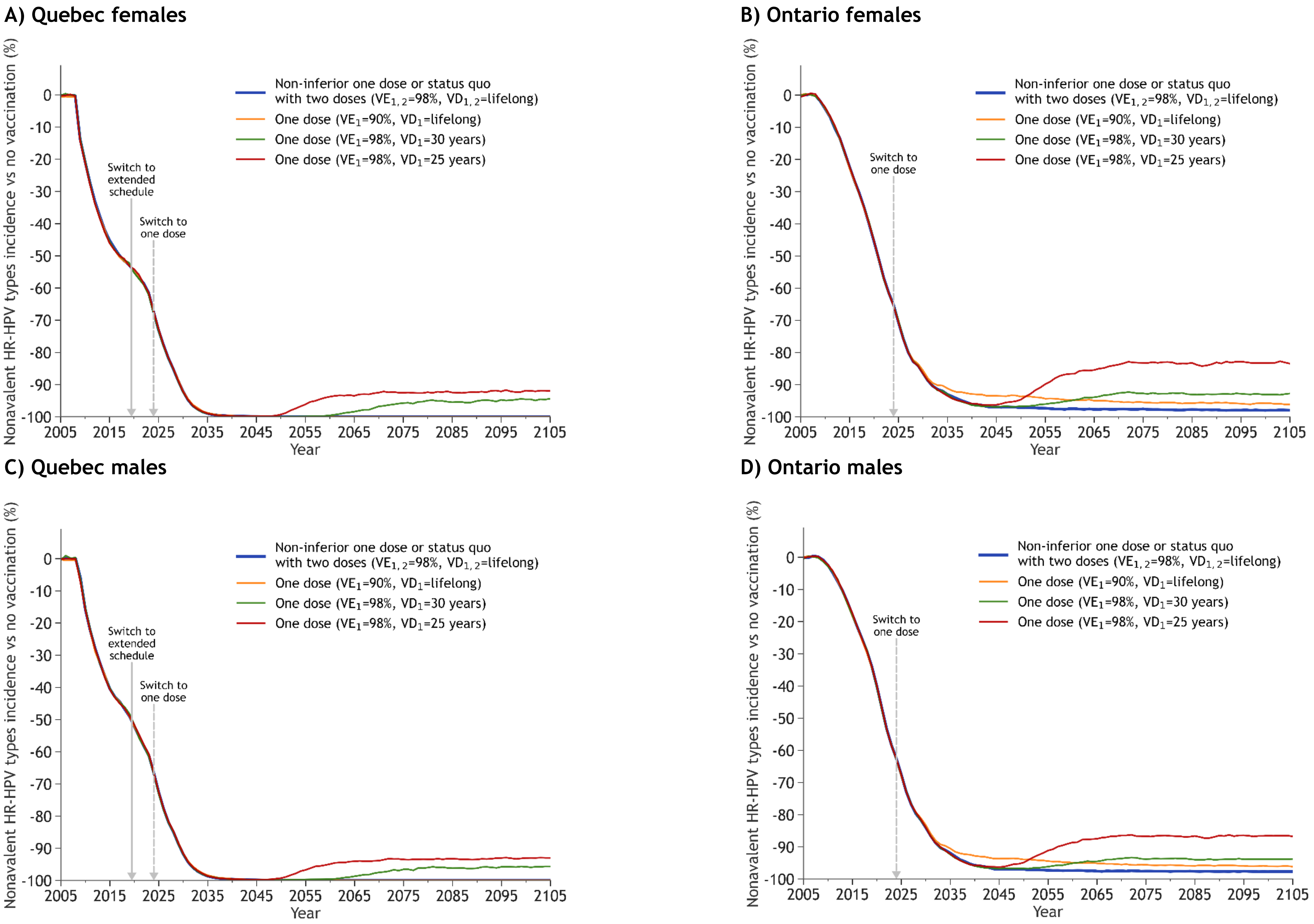

$VE_i$: Vaccine efficacy of dose *i*; $VD_i$: Vaccine duration of protection of dose *i*.

HR-HPV types: High-risk HPV types included in the nonavalent vaccine: 16, 18, 31, 33, 45, 52, 58

All panels: The lines are the median result of model projections using 50 parameter sets. All scenarios overlap during the first years after the start of vaccination.

Panels A,B: In Quebec the scenarios in blue and yellow lead to the elimination of HPV-16 and are hidden by the x axis. In 2019/2002, Quebec switched to a 5-year extended schedule with the 1st dose given at 9 years old and the 2nd dose to be given at 14 years old, from 2024/2025, if required. If the 2nd dose is not given, Quebec will have switched to one-dose schedule in 2019/2020.

**Figure A6. Projected population-level impact of switching to one-dose HPV vaccination for different one-dose efficacy and duration scenarios on the incidence of all other HPV-related cancers among females and males in Quebec and Ontario.**

**A) Quebec**

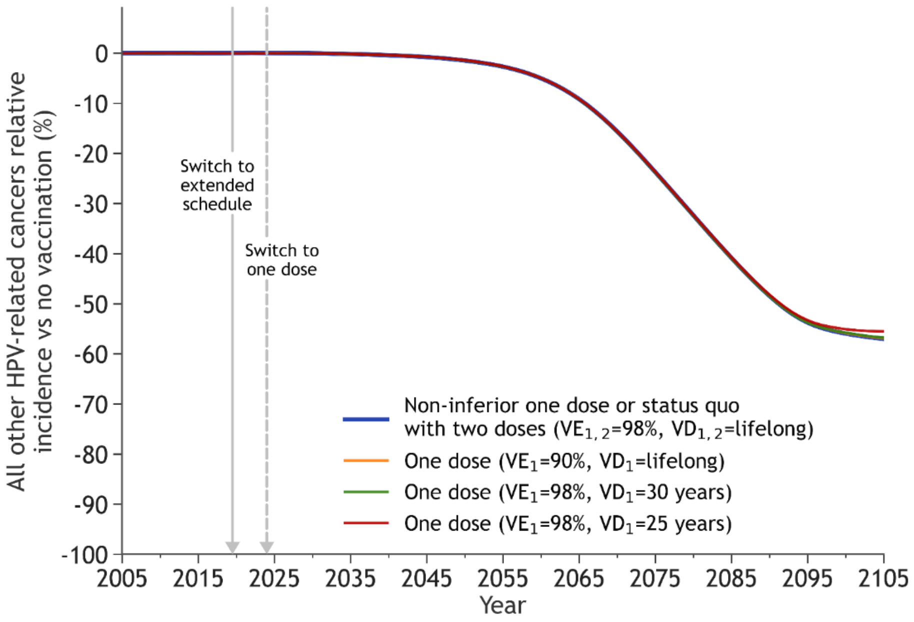


**B) Ontario**

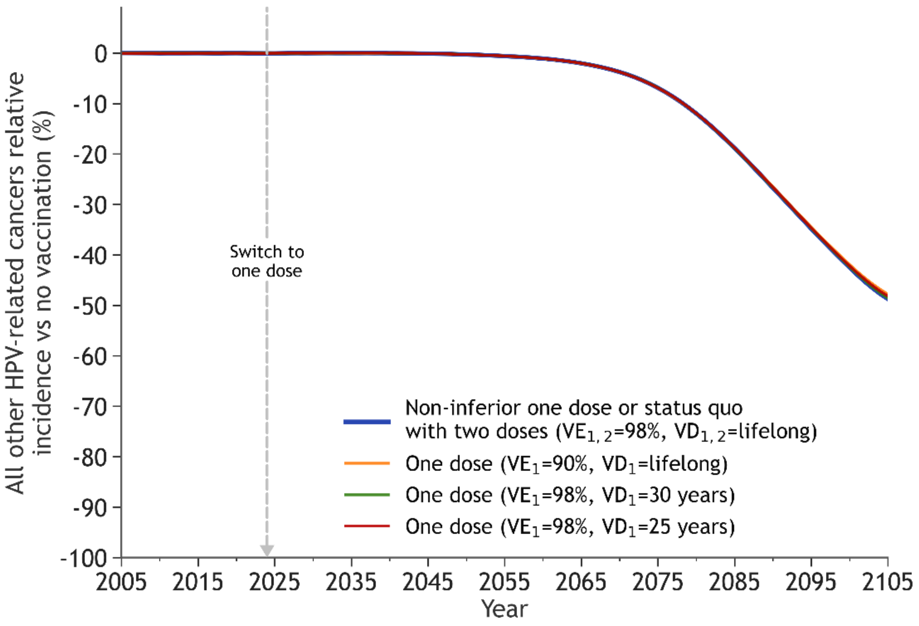


$VE_i$: Vaccine efficacy of dose *i*; $VD_i$: Vaccine duration of protection of dose *i*.

All panels: The lines are the median result of model projections using 50 parameter sets. All scenarios overlap during the first years after the start of vaccination.

Panel A: In 2019/2020, Quebec switched to a 5-year extended schedule with the 1st dose given at 9 years old and the 2nd dose to be given at 14 years old, from 2024/2025, if required. If the 2nd dose is not given, Quebec will have switched to one-dose schedule in 2019/2020.

**Figure A7. Sensitivity analysis – non-inferior one-dose for girls and worst-case one-dose for boys: projected population-level impact of switching to one-dose on HPV-16 infection and cervical cancer incidence among females and males from Quebec and Ontario.**

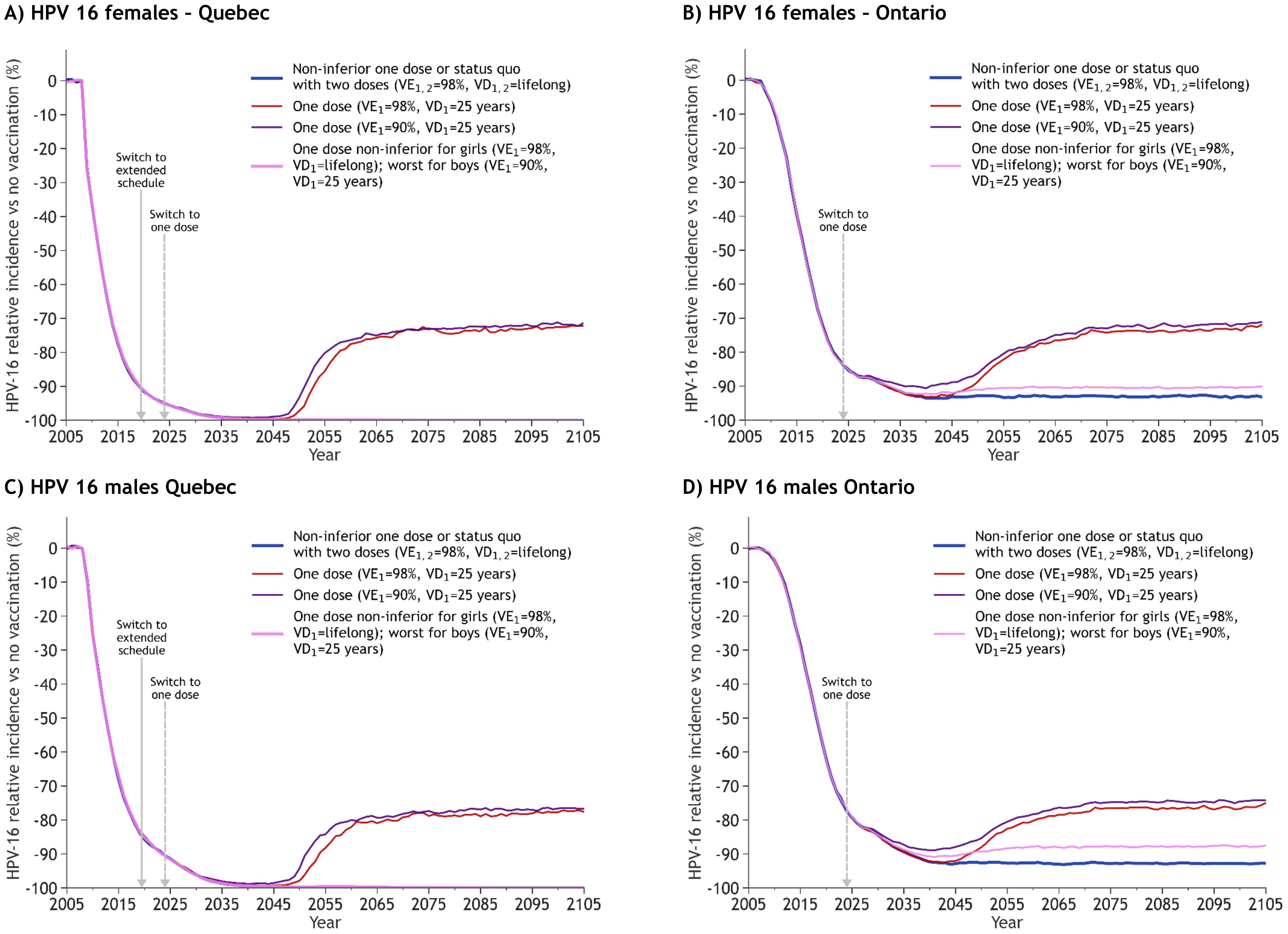

**E) Cervical cancer Quebec**

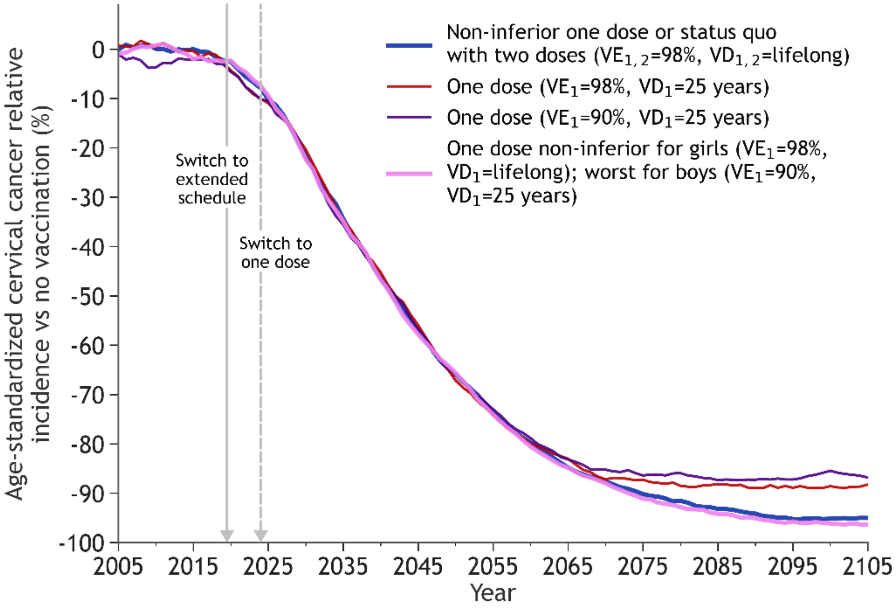


**F) Cervical cancer Ontario**

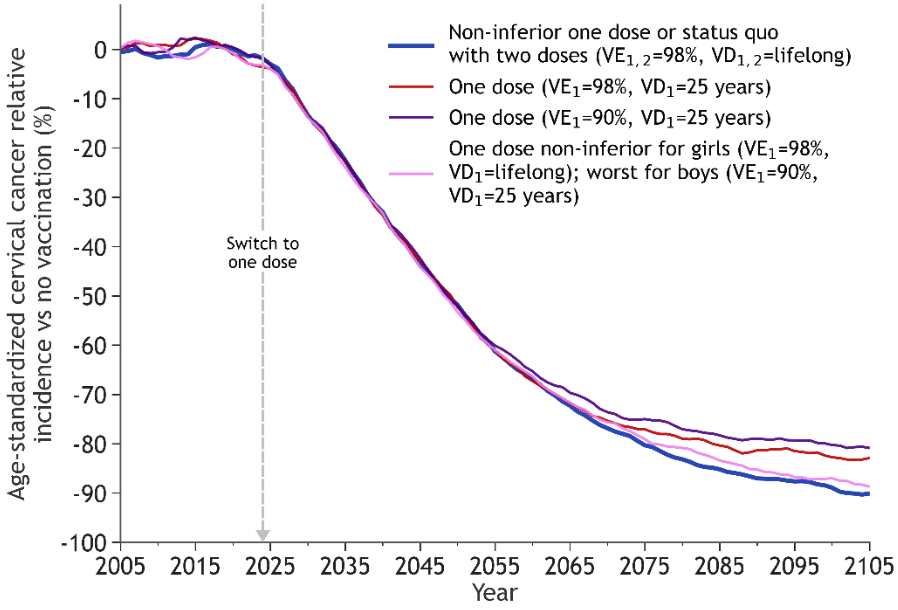


$VE_i$: Vaccine efficacy of dose *i*; $VD_i$: Vaccine duration of protection of dose *i*.

All panels: The lines are the median result of model projections using 50 parameter sets. All scenarios overlap during the first years after the start of vaccination.

Panels A,C,E: In 2019/2020, Quebec switched to a 5-year extended schedule with the 1st dose given at 9 years old and the 2nd dose to be given at 14 years old, from 2024/2025, if required. If the 2nd dose is not given, Quebec will have switched to one-dose schedule in 2019/2020.

**Figure A8. Sensitivity analysis – partial waning and lower sexual activity: projected population-level impact of switching to one-dose on HPV-16 infection and cervical cancer incidence among females and males from Quebec and Ontario.**

##### HPV-16 – Quebec females

A) Non-inferior one dose | B) 30 years one-dose duration | C) 25 years one-dose duration

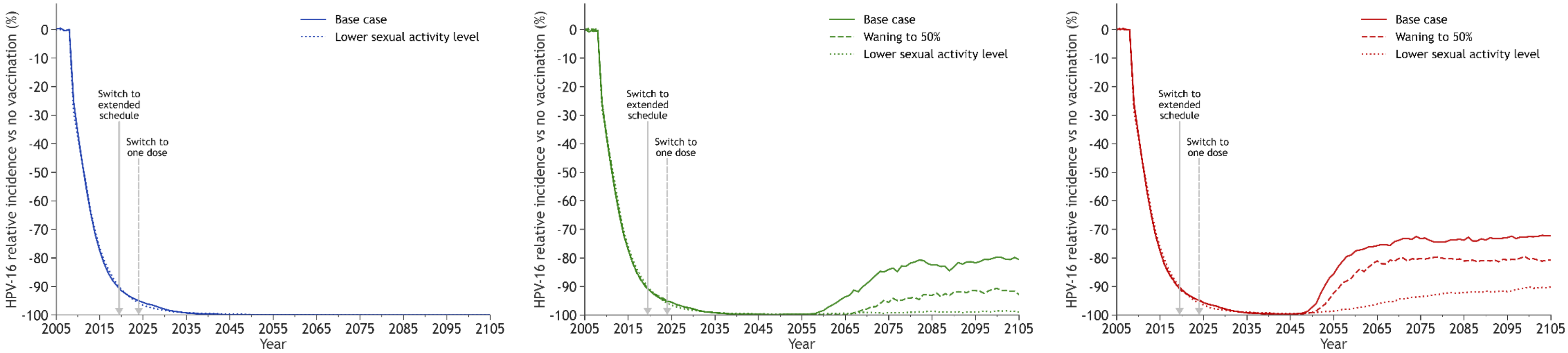


##### HPV-16 Ontario females

D) Non-inferior one dose | E) 30 years one-dose duration | F) 25 years one-dose duration

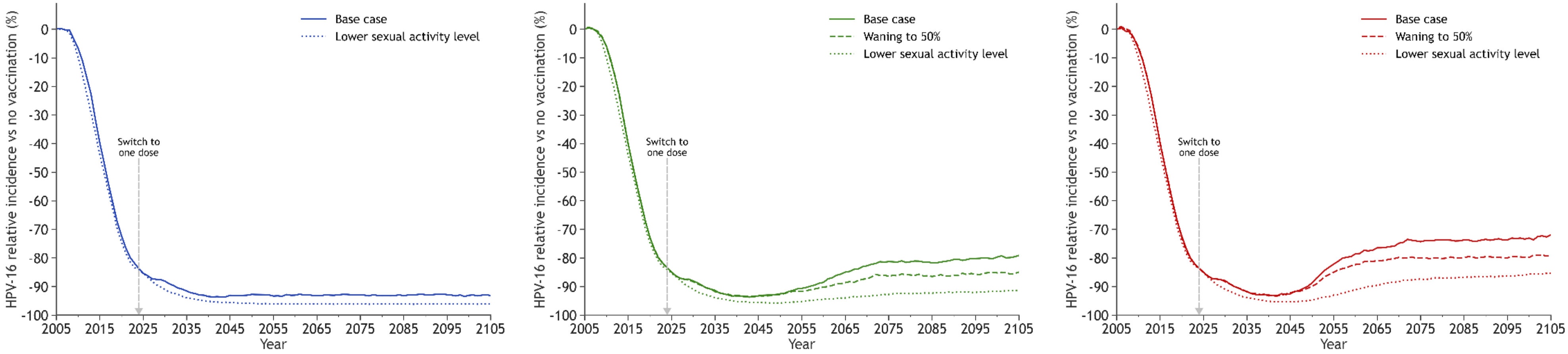

#### HPV-16 - Quebec males

##### G) Non-inferior one dose

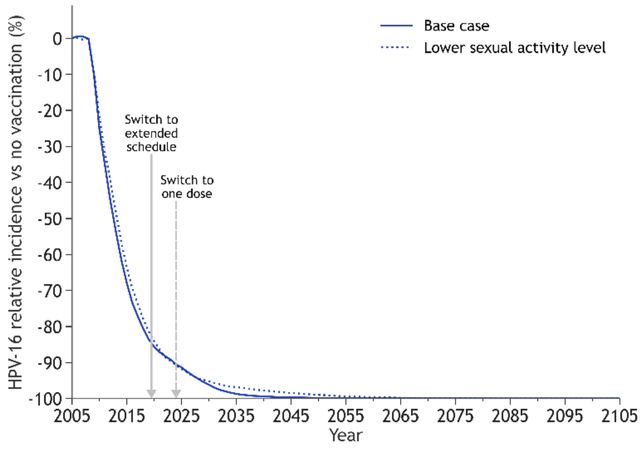


##### H) 30 years one-dose duration

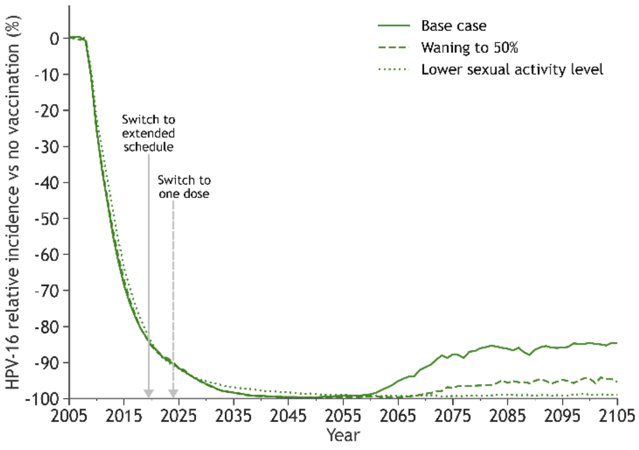


##### I) 25 years one-dose duration

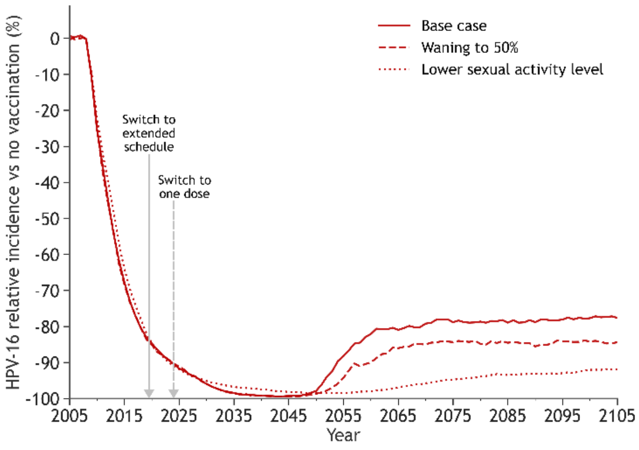


#### HPV-16 Ontario males

##### J) Non-inferior one dose

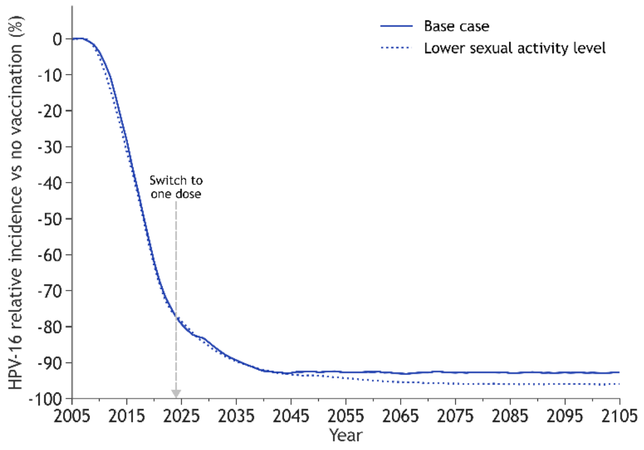


##### K) 30 years one-dose duration

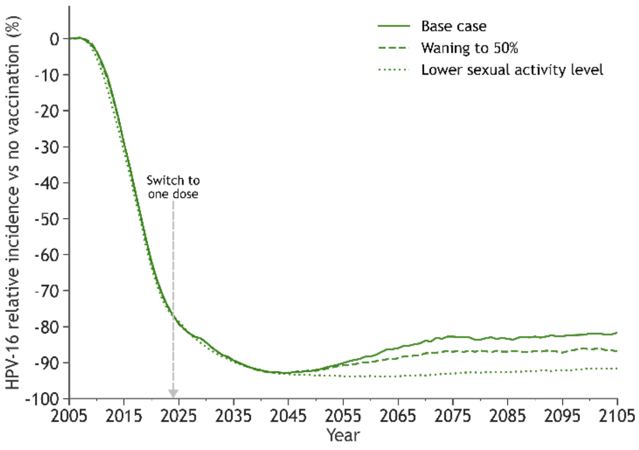


##### L) 25 years one-dose duration

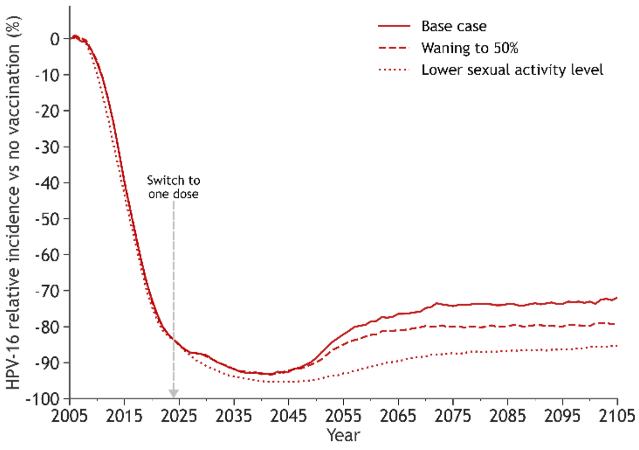

#### Cervical cancer – Quebec

##### M) Non-inferior one dose

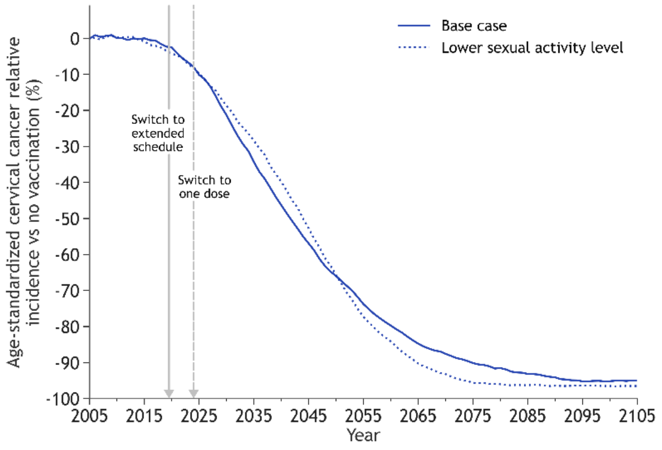


##### N) 30 years one-dose duration

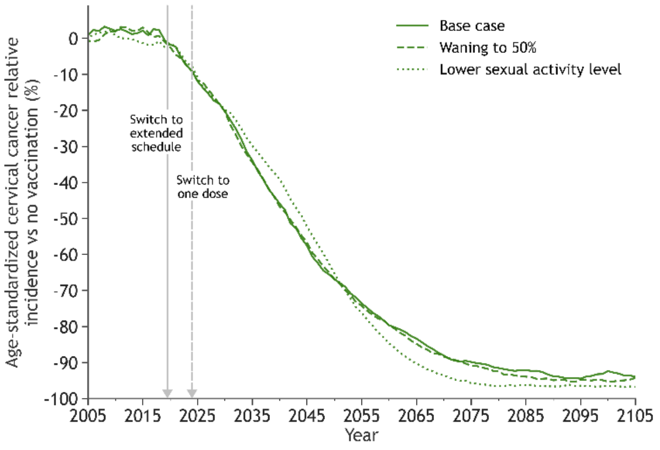


##### O) 25 years one-dose duration

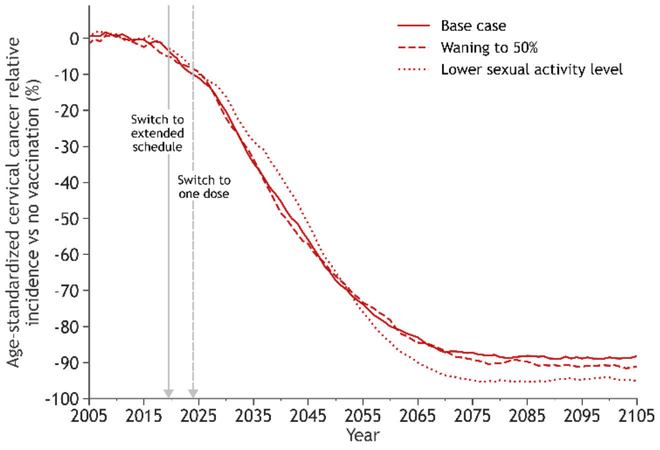


#### Cervical cancer – Ontario

##### P) Non-inferior one dose

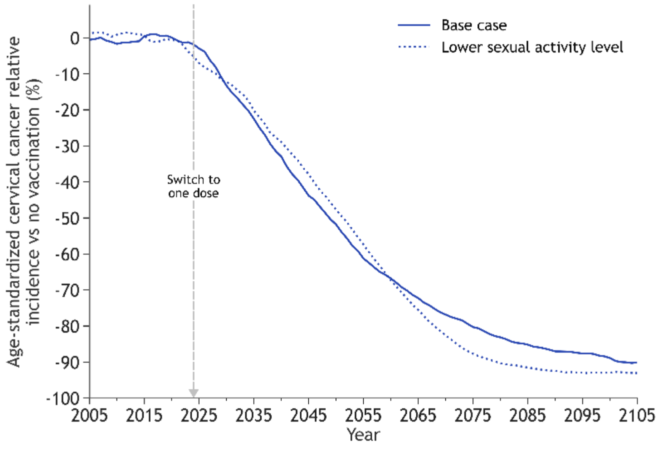


##### Q) 30 years one-dose duration

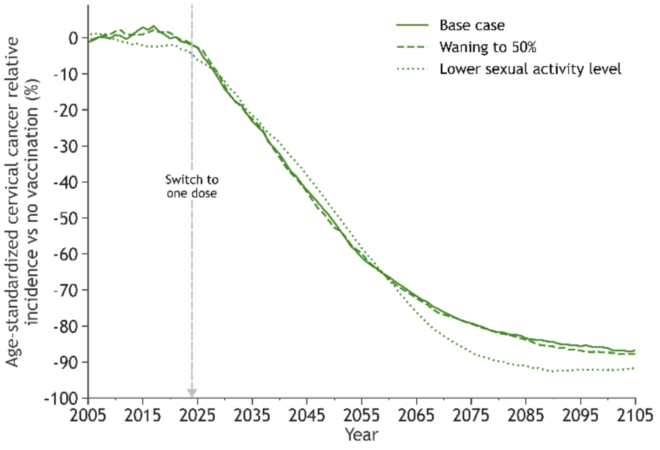


##### R) 25 years one-dose duration

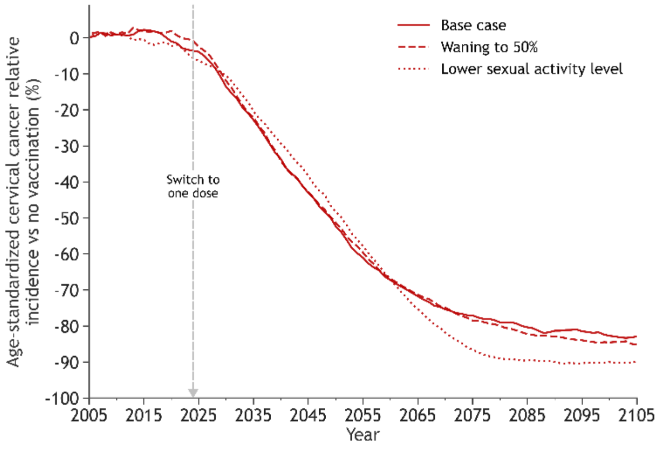

**Figure A9. Sensitivity analysis - partial waning and lower sexual activity: percent change in the cumulative number of cervical cancers averted over 100 years in Quebec and Ontario, for HPV vaccination with two doses and one dose, with different one-dose efficacy and duration scenarios.**

#### A) Quebec

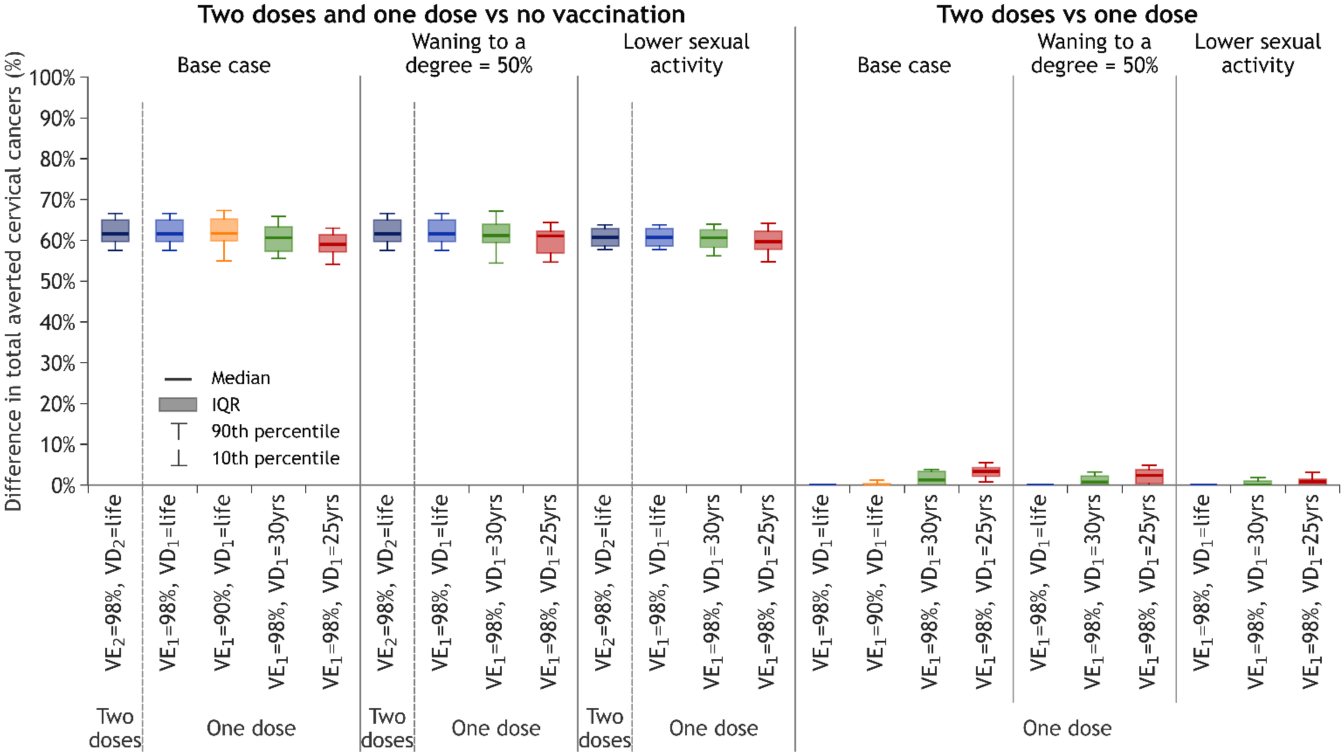


#### B) Ontario

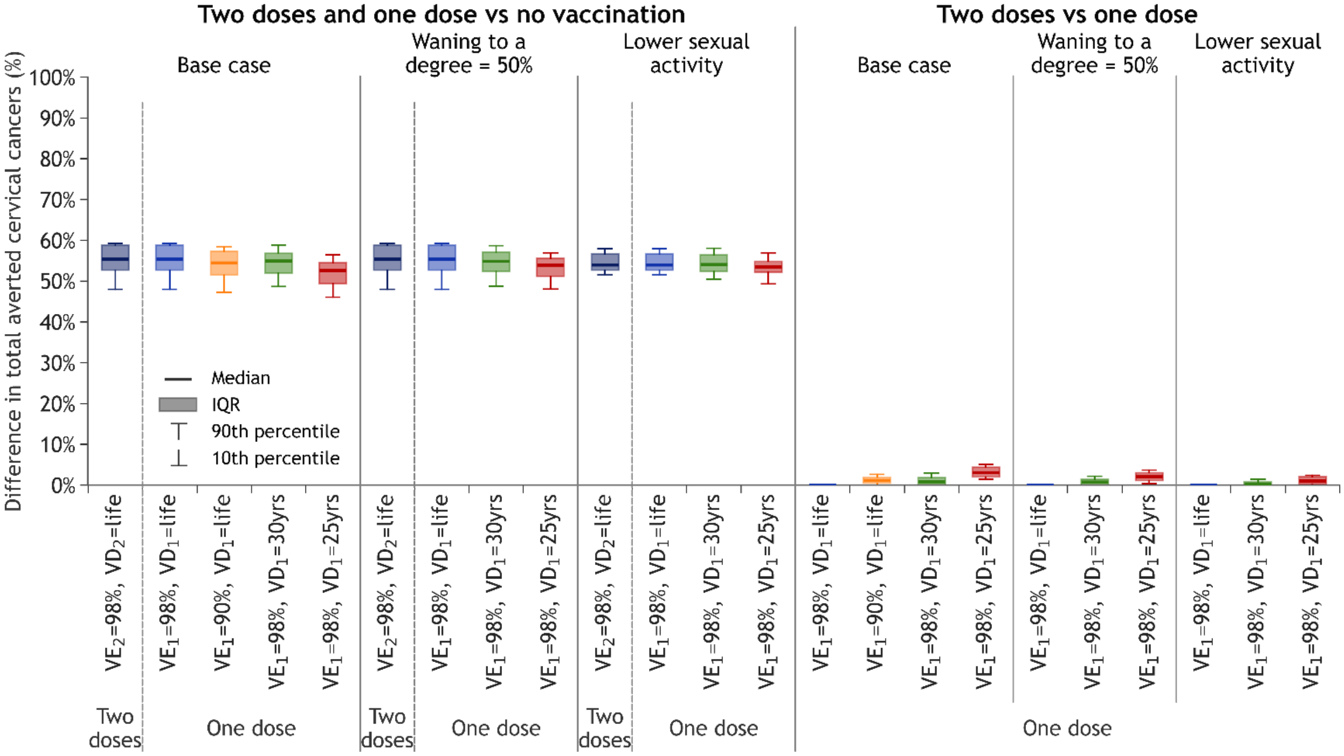

$VE_i$: Vaccine efficacy of dose $i$; $VD_i$: Vaccine duration of protection of dose $i$; IQR: Interquartile range.

At the left of each panel, two doses and one dose are compared to no vaccination; at the right of each panel, two doses are compared with one dose, for different one-dose efficacy and duration scenarios.

Boxplots represent the median, $10^{th}$, $25^{th}$, $75^{th}$ and $90^{th}$ percentiles of model projections using 50 parameter sets. This difference of averted cancer is estimated among all women of all ages since the beginning of vaccination and therefore includes unvaccinated and vaccinated cohorts.

**Figure A10. Sensitivity analysis - partial waning and lower sexual activity: number of doses needed to prevent one cervical cancer (NNV) in Quebec and Ontario for HPV vaccination with two doses and one dose, with different one-dose efficacy and duration scenarios.**

#### A) Quebec

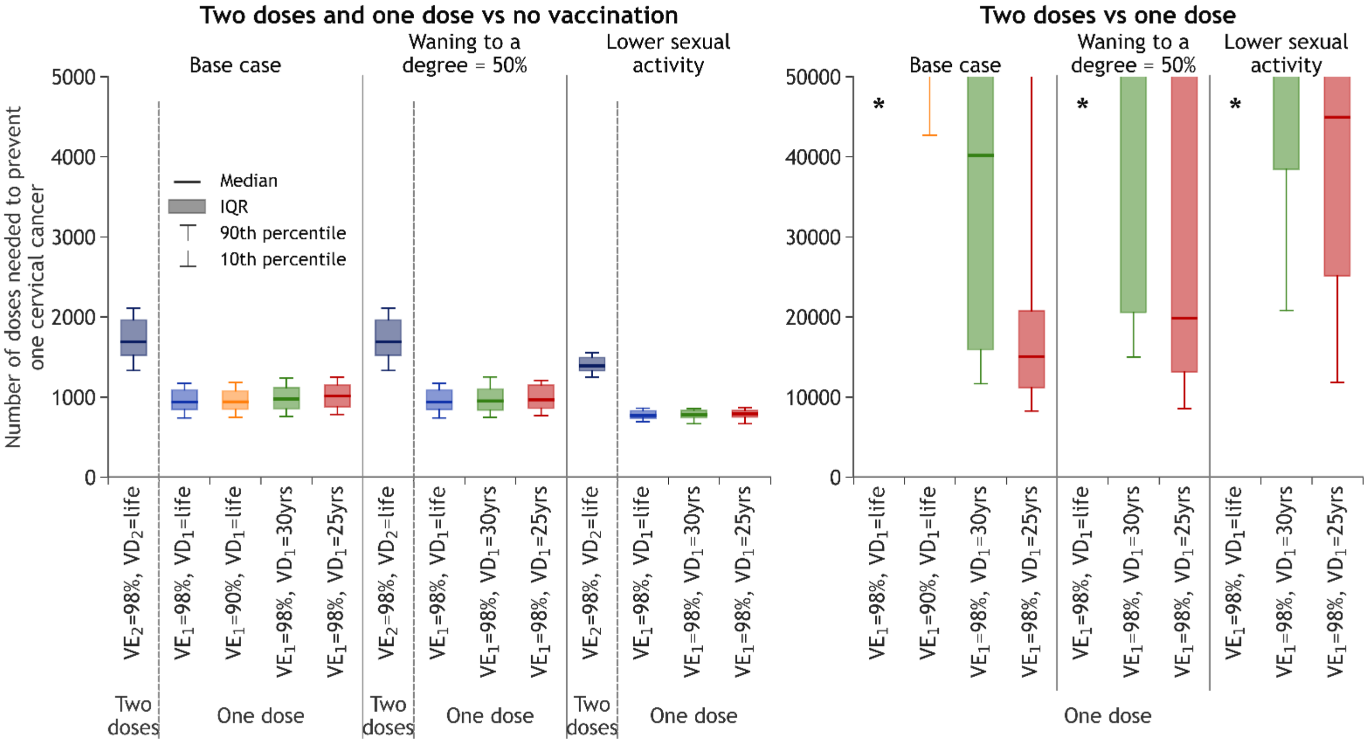


#### B) Ontario

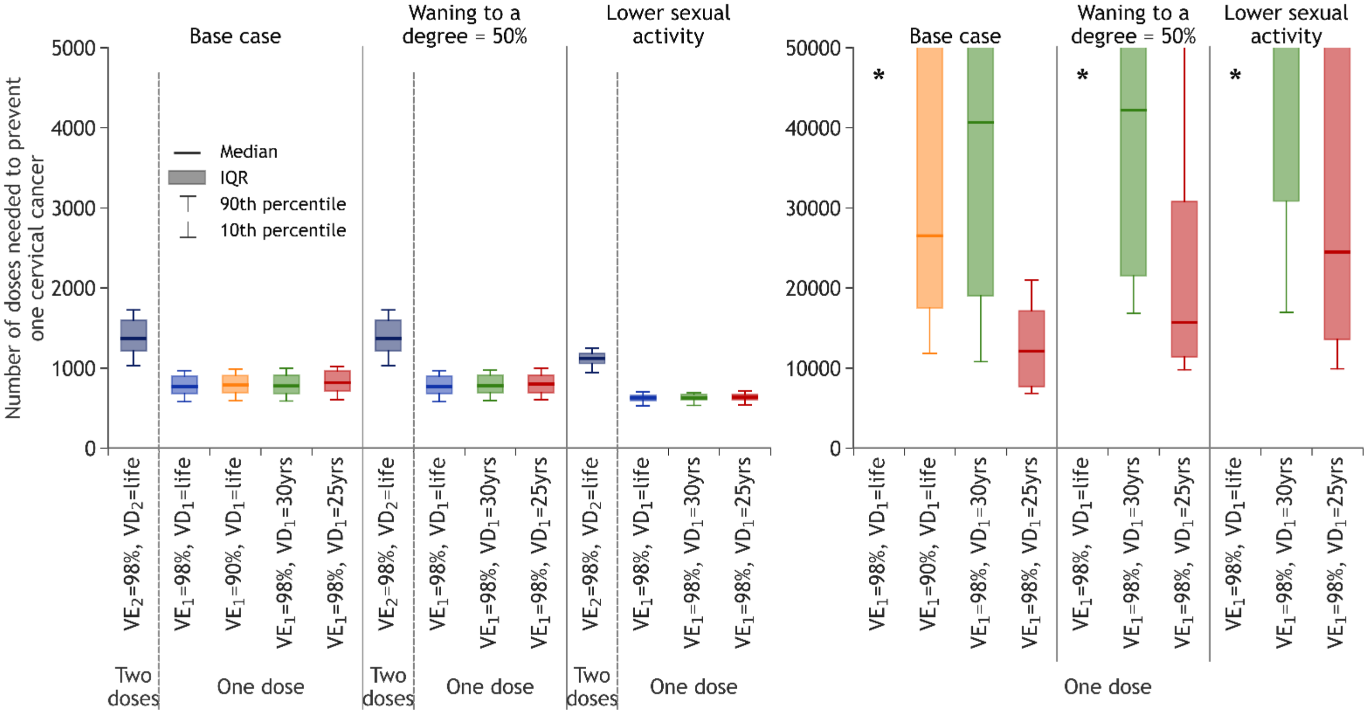

$VE_i$: Vaccine efficacy of dose *i*; $VD_i$: Vaccine duration of protection of dose *i*.

* Model projections of the incremental number of doses needed to prevent one cervical cancer are higher than 50,000 for this scenario.

At the left of each panel, two doses and one dose are compared to no vaccination; at the right of each panel, two doses are compared with one dose, for different one-dose efficacy and duration scenarios.

Boxplots represent the median, $10^{th}$, $25^{th}$, $75^{th}$ and $90^{th}$ percentiles of model projections using 50 parameter sets.

**Figure A11. Sensitivity analysis - mitigation strategy: projected population-level impact of switching back to two dose after 10 years of one-dose vaccination on HPV-16 infection and cervical cancer incidence among females and males from Quebec and Ontario.**

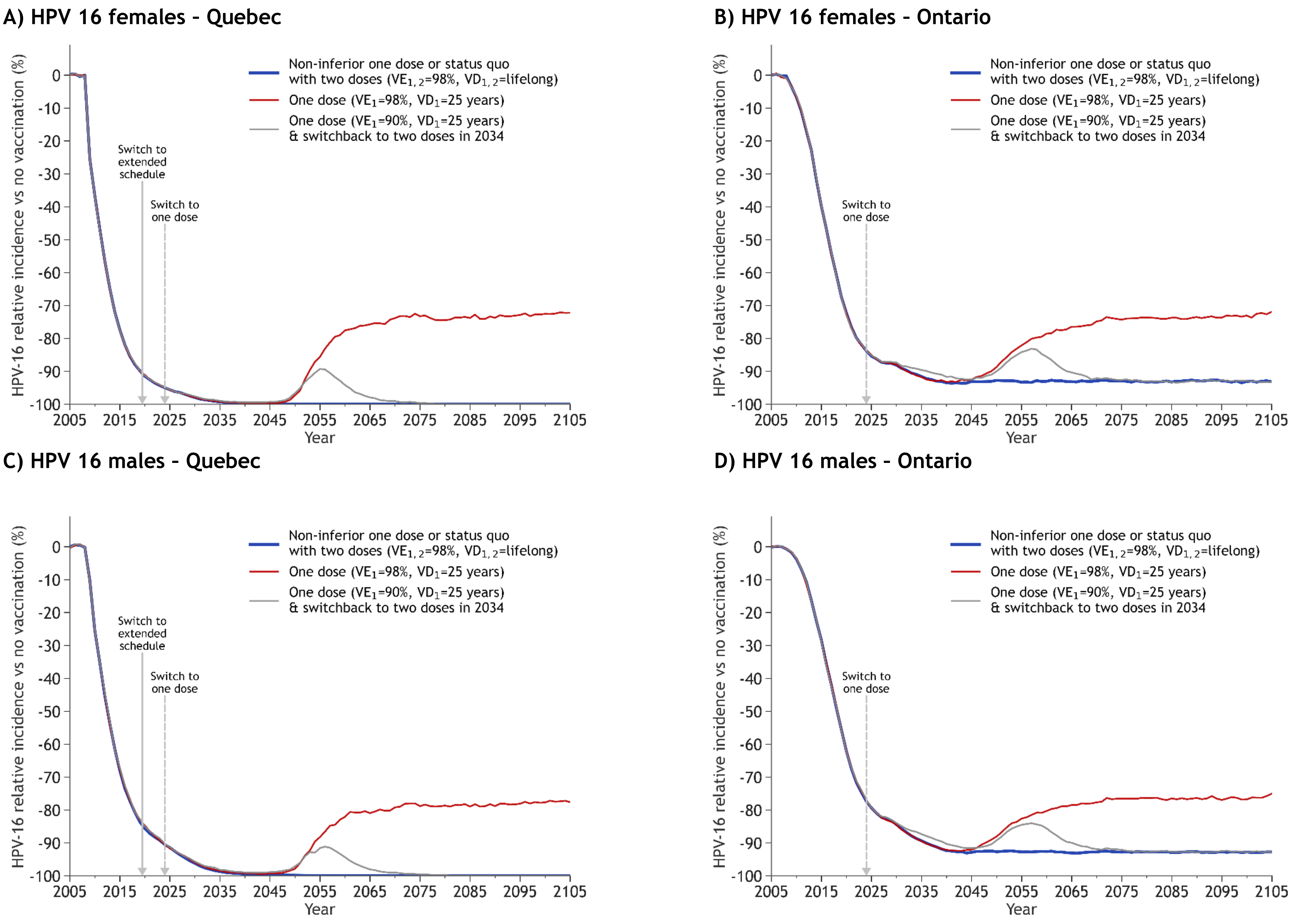

**E) Cervical cancer- Quebec**

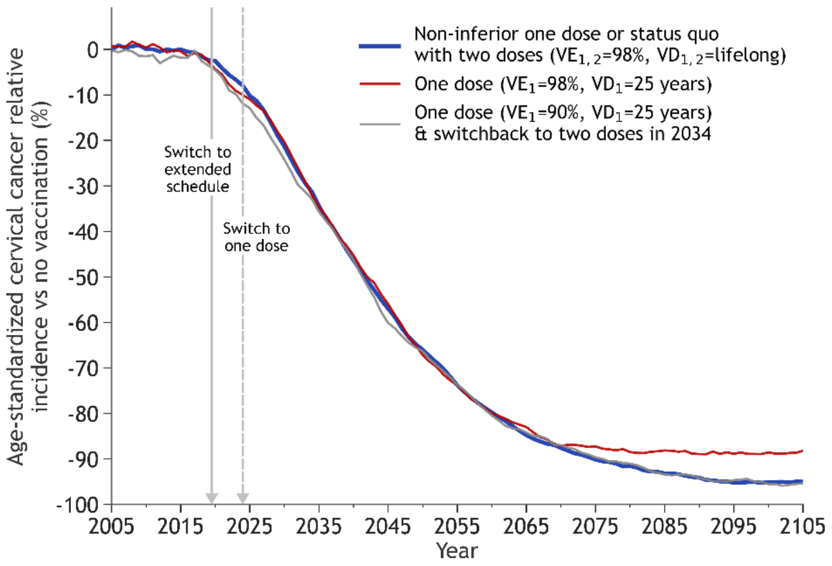


**F) Cervical cancer – Ontario**

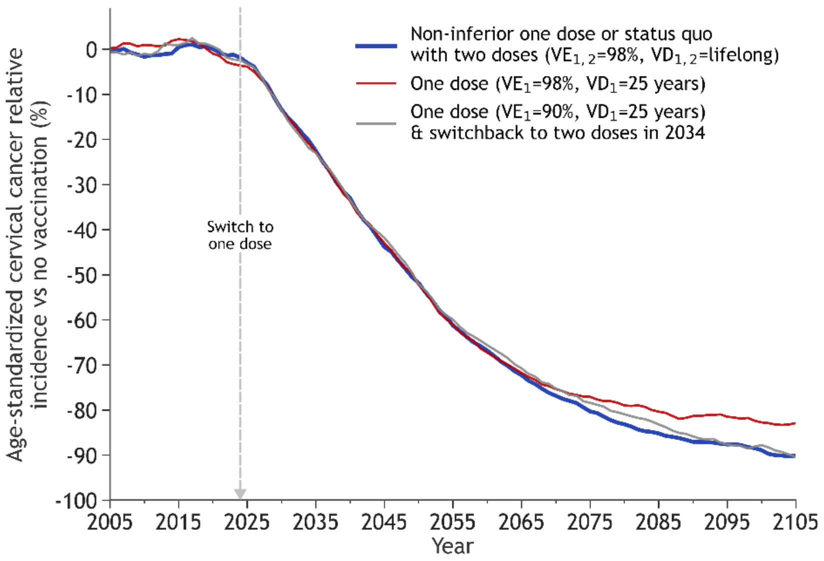


$VE_i$: Vaccine efficacy of dose *i*; $VD_i$: Vaccine duration of protection of dose *i*.

All panels: The lines are the median result of model projections using 50 parameter sets. All scenarios overlap during the first years after the start of vaccination.

Panels A,C,E: In 2019/2020, Quebec switched to a 5-year extended schedule with the 1st dose given at 9 years old and the 2nd dose to be given at 14 years old, from 2024/2025, if required. If the 2nd dose is not given, Quebec will have switched to one-dose schedule in 2019/2020.
